# School Closures due to Seasonal Influenza: Experience from Eleven Influenza Seasons – United States, 2011–2022

**DOI:** 10.1101/2023.08.28.23294732

**Authors:** Nicole Zviedrite, Ferdous Jahan, Yenlik Zheteyeva, Hongjiang Gao, Amra Uzicanin

## Abstract

While there are numerous studies about influenza pandemic-associated school closures, literature is scant regarding closures associated with seasonal influenza. To address this knowledge gap, we conducted systematic daily online searches from August 1, 2011– June 30, 2022, to identify public announcements of unplanned school closures in the US lasting ≥1 day, selecting those that mentioned influenza and influenza-like illness (ILI) as reason for school closure (ILI-SCs). We studied ILI-SC temporal patterns and compared them with reported outpatient ILI-related healthcare visits and laboratory confirmed influenza hospitalizations with attention to the difference between the pre-COVID-19 pandemic and the COVID-19-affected years. We documented that ILI-SCs occurred annually and concurrently with, and likely as a consequence of, widespread illness. The strongest correlations were primarily observed during influenza A (H3N2)-dominant seasons. ILI-SCs were heavily centered in HHS Region 4 and disproportionately impacted rural and lower-income communities.

**Article summary line:** Influenza-related school closures occurred annually in the US and their temporal patterns mirror the general patterns of influenza activity on both national and regional levels as observed through routine surveillance of medically attended ILI.

## Background

Because school-aged children and schools play important roles in accelerating community-wide influenza transmission, school closures implemented preemptively, i.e. before the virus is widespread, are an important nonpharmaceutical intervention (NPI) for severe influenza pandemics [1]. By contrast, influenza-associated closures that occur reactively, following already heightened levels of disease, are not considered an NPI [1].

We have previously reported, based on 2-year data (school years 2011/12 and 2012/13), that unplanned school closures (i.e., school closures not included in the academic calendar at the beginning of the school year) affect millions of students across the United States (US) annually and occur for various reasons, most frequently due to severe weather and natural disasters [2]. While illness-related closures constituted only about 1% of all unplanned school closures, almost 60% of them were related to respiratory illnesses and their annual occurrence pattern mirrored national influenza activity [2].

In the present study, we describe the multi-year pattern of influenza-like illness (ILI)-related school closures in the US over eleven consecutive school years, 2011/12 through 2021/22, with attention to the difference between the pre-COVID-19 pandemic and the COVID-19-affected years, and analyze geographic and temporal relationships between ILI-related school closures and routine surveillance data on medically-attended influenza and ILI on national and regional levels.

## Methods

### Data collection

From August 1, 2011, through June 30, 2022, we conducted systematic daily searches of publicly available online data (via Google and Lexis-Nexis) to identify unplanned kindergarten through twelfth grade (K–12) school closure events in the US using search strategy and data abstraction methodology described in detail by Wong et al. [2]. Closure events were documented at either the district or individual school level, based on the scope of the closure decision as reported in the announcements. For present analysis, we selected school closure events for which influenza- and/or ILI (ILI-SCs) were mentioned in the public announcements as a reason for the closure decision. School closures attributed to Coronavirus Disease 2019 (COVID-19), in absence of influenza, were excluded. From each ILI-SC announcement, we abstracted contributing factors specific to ILI and grouped them into five non-mutually exclusive categories as seen in Table S1.

We downloaded publicly available data on school and school district characteristics from the National Center for Education Statistics’ (NCES) Common Core of Data, which includes all primary and secondary public schools and districts, and Private School Universe Survey, which has reported a high coverage rate of traditional private schools (ranging from 89.6% - 98.8% from 2003/04 – 2015/16). Datapoints included number of schools in affected school districts, number of enrolled students, and number of staff [3, 4], which were linked with ILI-SC data by NCES school district or individual school identifiers. For school district-level ILI-SCs, we further disaggregated districts into their individual schools. NCES data for the year when the ILI-SC occurred were used, when possible, or, otherwise, for the most recent year of data available. Districts with zero schools; schools with zero students; vocational, special education, and alternative schools with missing value for students; online/virtual schools; adult education programs; summer schools; jails; and schools with pre-kindergarten or transitional kindergarten as the highest grade were excluded from analysis.

We gathered publicly available surveillance data on seasonal influenza activity, including weekly national and US Department of Health and Human Services (HHS) region-specific [5] data reporting percent of outpatient medical provider visits for influenza-like illness (ILINet) [6] and national laboratory-confirmed influenza-associated hospitalizations of children and adults (FluSurv-NET) reported by thirteen states (California, Colorado, Connecticut, Georgia, Maryland [Baltimore metropolitan area], Michigan, Minnesota, New Mexico, New York, Ohio, Oregon, Tennessee, and Utah), [7] from August 1, 2011–June 30, 2022, as well as reported seasonal severity and predominant strain(s) [8].

### Data management and analysis

Data were imported into SAS 9.4 (SAS Institute Inc., Cary, North Carolina) for analysis. Descriptive statistics were calculated to summarize characteristics of ILI-SCs (cause, seasonality, duration, and geographic distribution) and impacts on closed schools (number of students and teachers affected).

We studied temporal patterns of weekly occurrence of ILI-SCs and compared them with seasonal influenza surveillance data. We calculated Spearman rank correlations to evaluate these relationships (at alpha=0.05) during influenza seasons (defined as epidemiological weeks 40 through 20 (ILINet), or through week 17 (FluSurv-NET)). P-value calculation for the Spearman’s rank correlation was based on Fisher’s Z-transformation. The final week of each calendar year (epidemiological weeks 52 (2011–2013, 2015–2019, 2021) or 53 (2014, 2020)) was excluded from analysis based on the assumption that all schools in the nation are on winter break during that period. In the 2019/20 season, we truncated the season after epidemiological week 11 of 2020 (March 8-14, 2020), which was the last week an ILI-SC was captured by our searches. Despite this, data from January and February 2020 may have been impacted by unrecognized transmission of severe acute respiratory syndrome coronavirus 2 (SARS-CoV-2), the causative agent of COVID-19 [9]. Truncation coincides with nationwide school closures implemented by states in response to the known spread of SARS-CoV-2 [10].

Additional analyses are described in supplementary material.

## Results

From August 1, 2011–June 30, 2022, we found 2,077 school closure events attributed to influenza/ILI; the average number of ILI-SC events per year during the pre-COVID-19 period (2011/12 to 2019/20) was 224·3 (median 112), while the average for the COVID-19-affected period (2020/21 and 2021/22) was 29 (5 and 53 events, respectively) (Table 1). In both the pre-COVID-19 and COVID-19-affected periods, the most frequently specified reason for closure was increased absenteeism of students and staff due to illness [1,358 (67·3%) and 34 (58·6%), respectively] (Table 2).

**Table 1.**
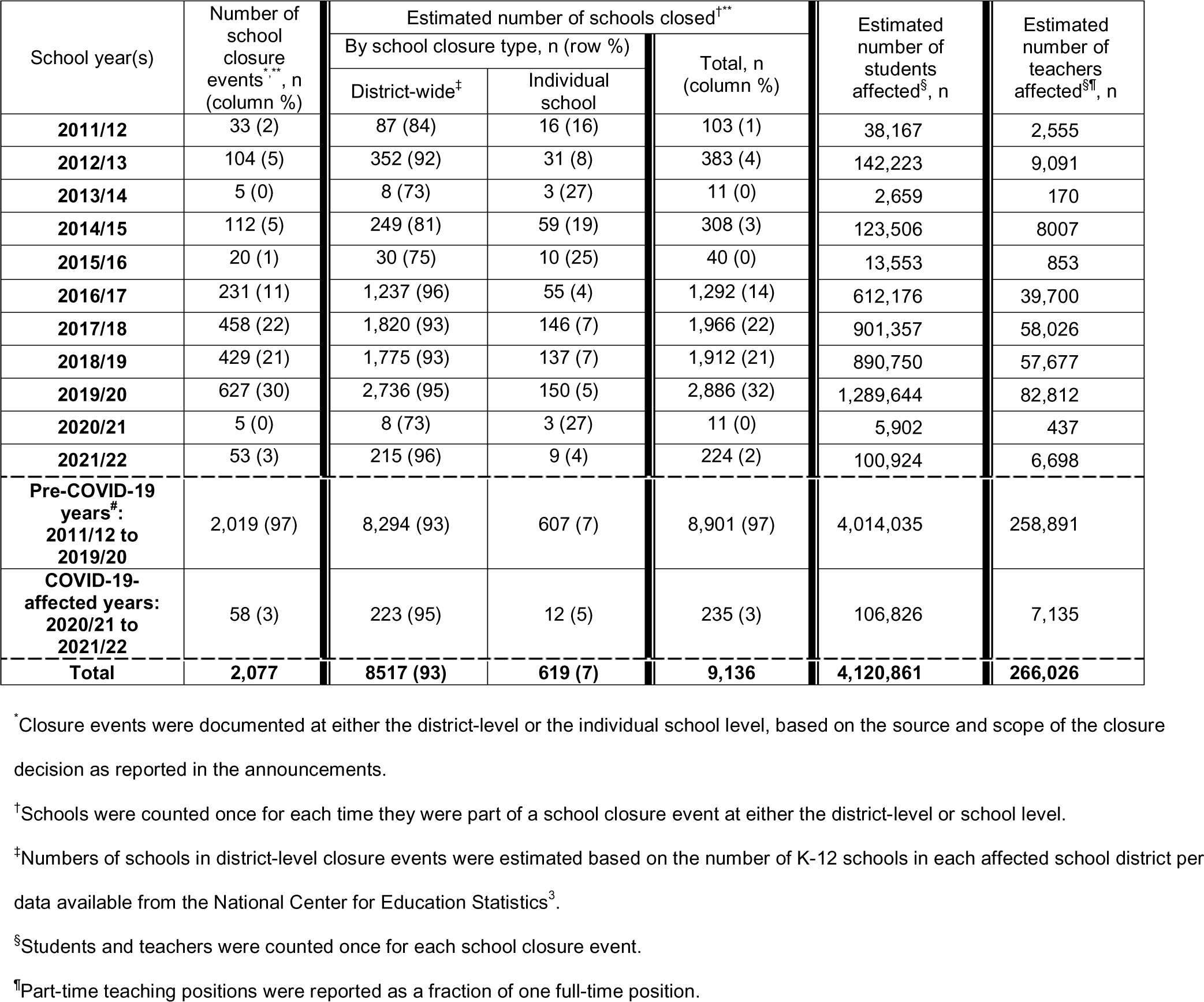
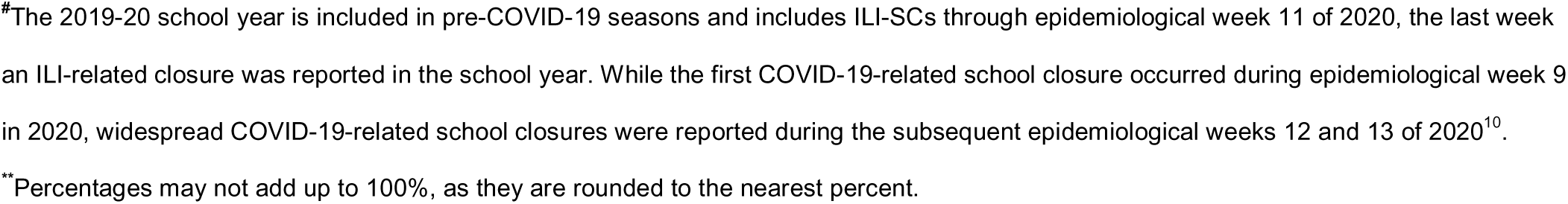
Publicly announced school closures associated with influenza-like illness, by school year – United States, August 1, 2011—June 30, 2022.

| School year(s) | Number of school closure events<sup>†,††</sup>, n (column %) | Estimated number of schools closed<sup>†**</sup> |  |  | Estimated number of students affected<sup>§</sup>, n | Estimated number of teachers affected<sup>§¶</sup>, n |
| --- | --- | --- | --- | --- | --- | --- |
|  |  | By school closure type, n (row %) |  | Total, n (column %) |  |  |
|  |  | District-wide<sup>‡</sup> | Individual school |  |  |  |
| 2011/12 | 33 (2) | 87 (84) | 16 (16) | 103 (1) | 38,167 | 2,555 |
| 2012/13 | 104 (5) | 352 (92) | 31 (8) | 383 (4) | 142,223 | 9,091 |
| 2013/14 | 5 (0) | 8 (73) | 3 (27) | 11 (0) | 2,659 | 170 |
| 2014/15 | 112 (5) | 249 (81) | 59 (19) | 308 (3) | 123,506 | 8007 |
| 2015/16 | 20 (1) | 30 (75) | 10 (25) | 40 (0) | 13,553 | 853 |
| 2016/17 | 231 (11) | 1,237 (96) | 55 (4) | 1,292 (14) | 612,176 | 39,700 |
| 2017/18 | 458 (22) | 1,820 (93) | 146 (7) | 1,966 (22) | 901,357 | 58,026 |
| 2018/19 | 429 (21) | 1,775 (93) | 137 (7) | 1,912 (21) | 890,750 | 57,677 |
| 2019/20 | 627 (30) | 2,736 (95) | 150 (5) | 2,886 (32) | 1,289,644 | 82,812 |
| 2020/21 | 5 (0) | 8 (73) | 3 (27) | 11 (0) | 5,902 | 437 |
| 2021/22 | 53 (3) | 215 (96) | 9 (4) | 224 (2) | 100,924 | 6,698 |
| Pre-COVID-19 years<sup>‡</sup>: 2011/12 to 2019/20 | 2,019 (97) | 8,294 (93) | 607 (7) | 8,901 (97) | 4,014,035 | 258,891 |
| COVID-19-affected years: 2020/21 to 2021/22 | 58 (3) | 223 (95) | 12 (5) | 235 (3) | 106,826 | 7,135 |
| Total | 2,077 | 8517 (93) | 619 (7) | 9,136 | 4,120,861 | 266,026 |

<sup>\*</sup>Closure events were documented at either the district-level or the individual school level, based on the source and scope of the closure decision as reported in the announcements.
<sup>†</sup>Schools were counted once for each time they were part of a school closure event at either the district-level or school level.
<sup>‡</sup>Numbers of schools in district-level closure events were estimated based on the number of K-12 schools in each affected school district per data available from the National Center for Education Statistics<sup>3</sup>.
<sup>§</sup>Students and teachers were counted once for each school closure event.
<sup>††</sup>Part-time teaching positions were reported as a fraction of one full-time position.
\*The 2019-20 school year is included in pre-COVID-19 seasons and includes ILI-SCs through epidemiological week 11 of 2020, the last week an ILI-related closure was reported in the school year. While the first COVID-19-related school closure occurred during epidemiological week 9 in 2020, widespread COVID-19-related school closures were reported during the subsequent epidemiological weeks 12 and 13 of 2020<sup>10</sup>.
\*\*Percentages may not add up to 100%, as they are rounded to the nearest percent.

**Table 2.**
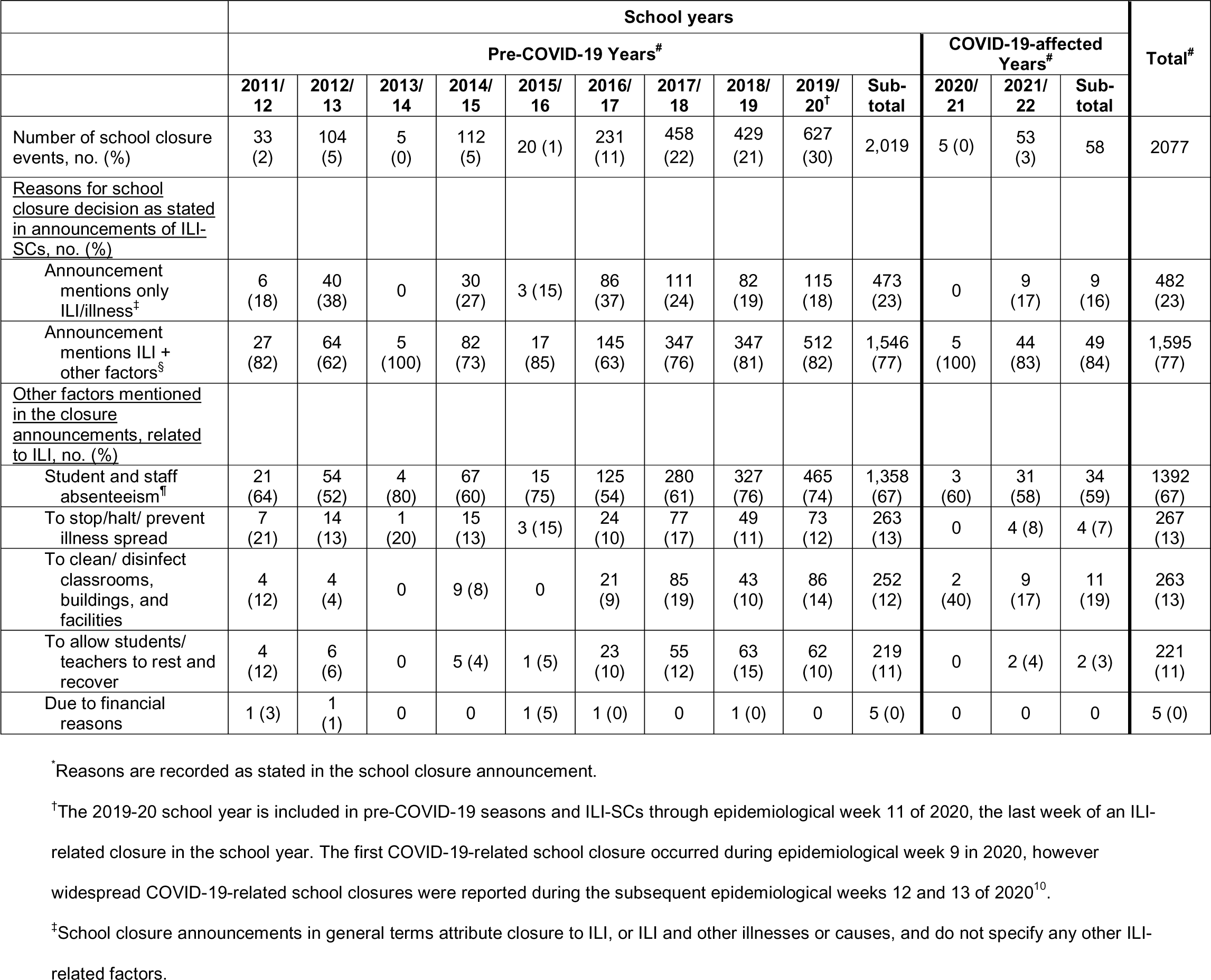
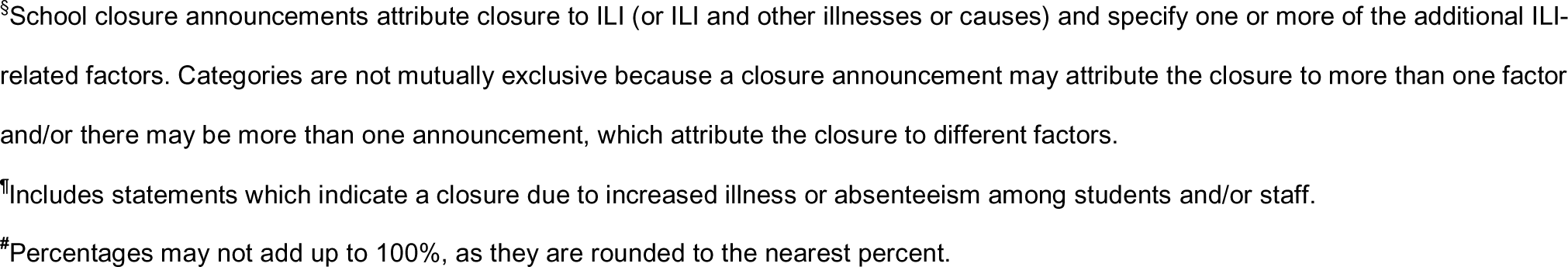
Reasons* for influenza-like illness-related school closure events, by school year – United States, August 1, 2011—June 30, 2022.

Over the 11-year study period, the 2,077 ILI-SC events accounted for an estimated 9,136 school closures and affected an estimated 4.1 million students and 260,000 teachers (Table 1, Fig 1). Nearly two-thirds of all ILI-SCs [6,040 (66·1%)] occurred in HHS Region 4, one of two regions to have at least one ILI-SC announced in every year of the study, the other being HHS Region 5 which accounted for one-tenth of ILI-SCs [991 (10·8%)] (Table S2). By contrast, ILI-SCs were seldom reported in some of the other regions. For example, in Region 1 twenty-one ILI-SCs were identified and in Region 9 only fourteen ILI-SCs were identified during the study period (Fig 2, Table S2). Correlation between ILI-SCs and ILINet data varied by HHS Region. Across eleven school years, moderate correlation between ILI-SCs and outpatient ILI surveillance was observed in Region 4 [r_s_=0·66 (p<0·001)], Region 5 [r_s_=0·61 (p<0·001), and Region 6 [r_s_=0·75 (p<0·001)] (Fig 2, Table S3). In other regions the correlation was weak or very weak.

**Fig 1.**
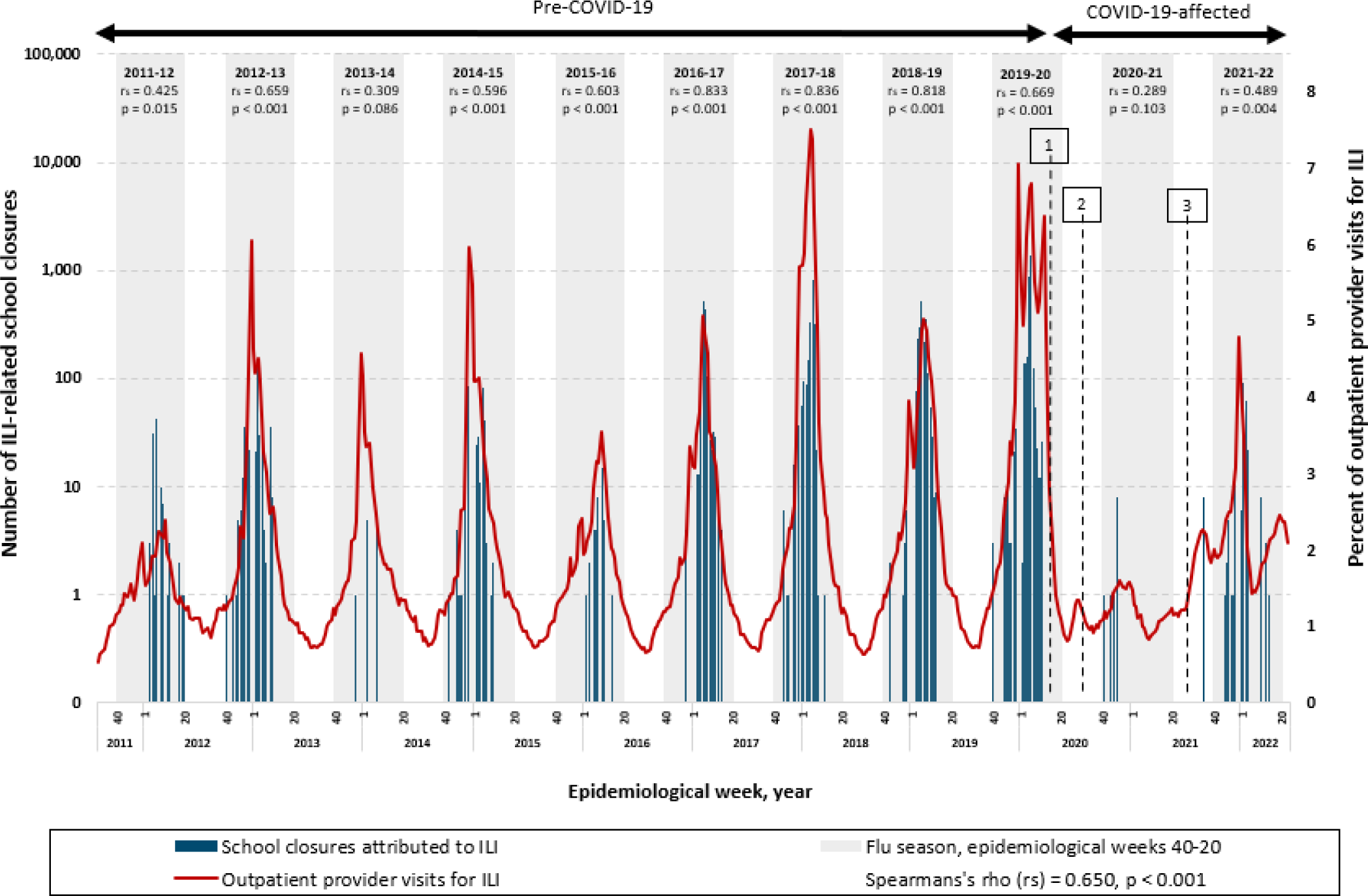
Estimated number of school closures associated with influenza-like illness (ILI) (N=9,136) and percent of outpatient provider visits for ILI* by epidemiologic week – United States, August 1, 2011—June 30, 2022. * Percent of outpatient provider visits for ILI per data available from ILINet [6]. 1) Schools closed nationwide due to spread of SARS COV2 [10]. 2) The majority of the schools opened virtually for the 2020-2021 SY, with in-person learning increasing over the school year [17]. 3) Schools reopened for the 2021-2022 SY with the majority in person [19]. Note: In the 2012-13, 2014-15, 2016-17, and 2017-18 influenza seasons, influenza A (H3N2) was the predominant strain [8]. In the 2011-12 influenza season, which was unusually mild, influenza A (H3N2) predominated overall but, influenza A (H1N1)pdm09 and influenza B also widely circulated [8]. In the 2013-14 and 2015-16 seasons, influenza A (H1N1)pdm09 was the predominant strain [8]. In the 2018-19 season, there were two peaks of similar magnitude dominated by influenza A (H1N1) followed by influenza A (H3N2) [8]. During the 2019-2020 season, Influenza B predominated early in the season followed by influenza A (H1N1)pdm09 [8]. In the 2020-2021 season, there was unusual low flu activity in the United States when both Influenza A ((H1N1)pdm09)and (H3N2)) and influenza B, and the majority of influenza A viruses were H3N2 [8]. In the 2021-2022 season, the majority of positive flu tests reported to the CDC by US Public Health Laboratories were attributed to influenza A (H3N2) [8]

**Fig 2.**
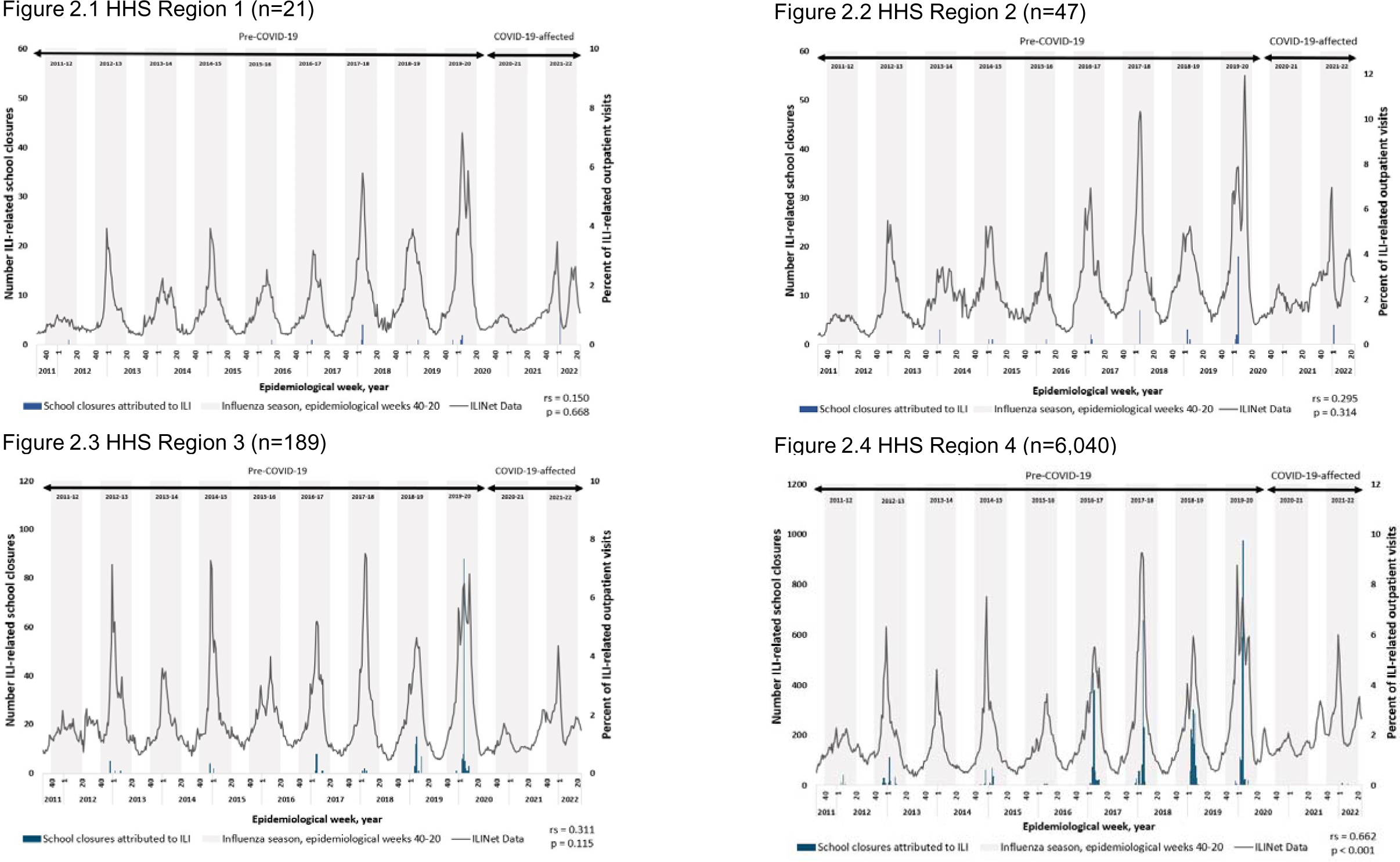
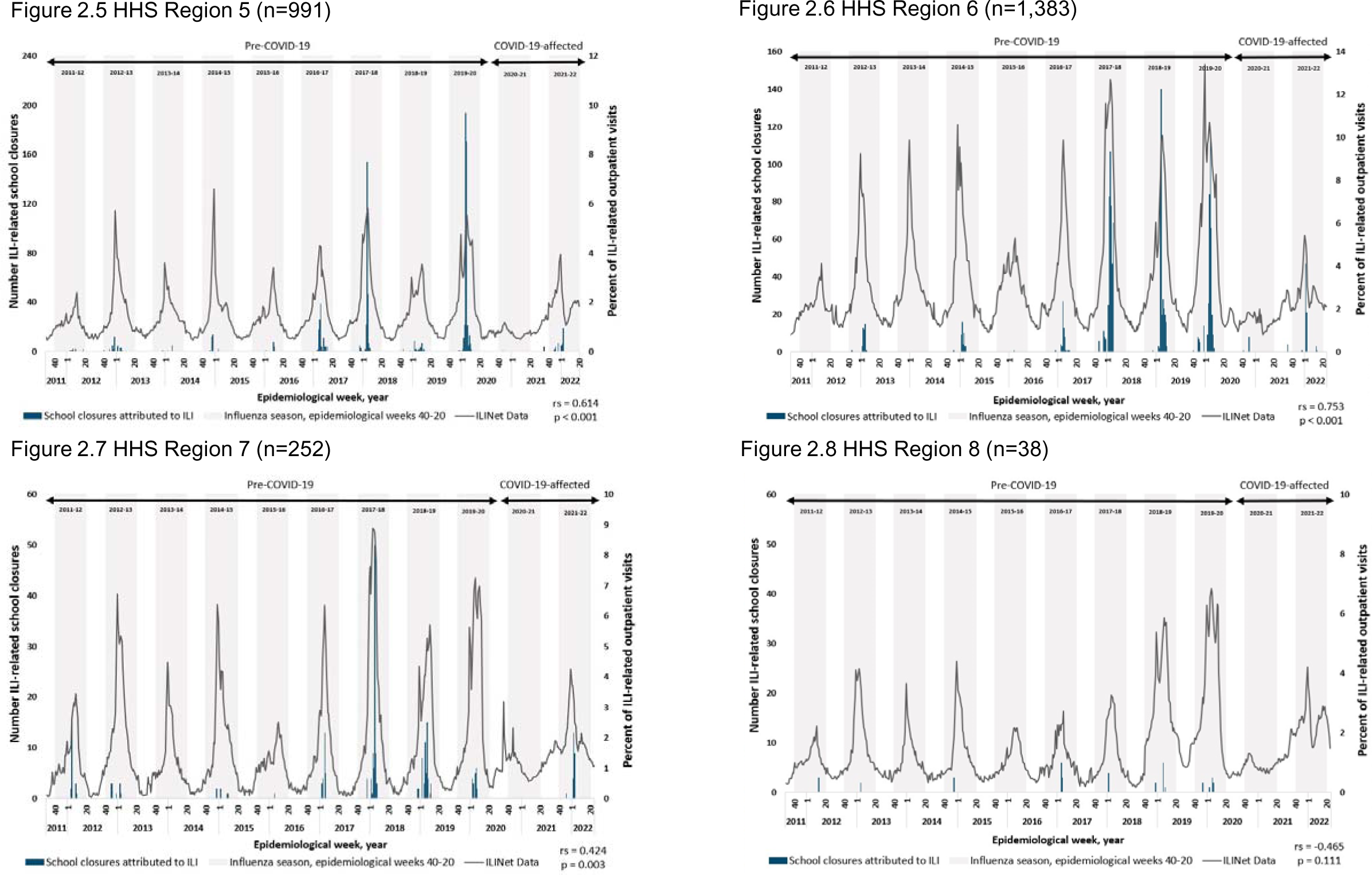
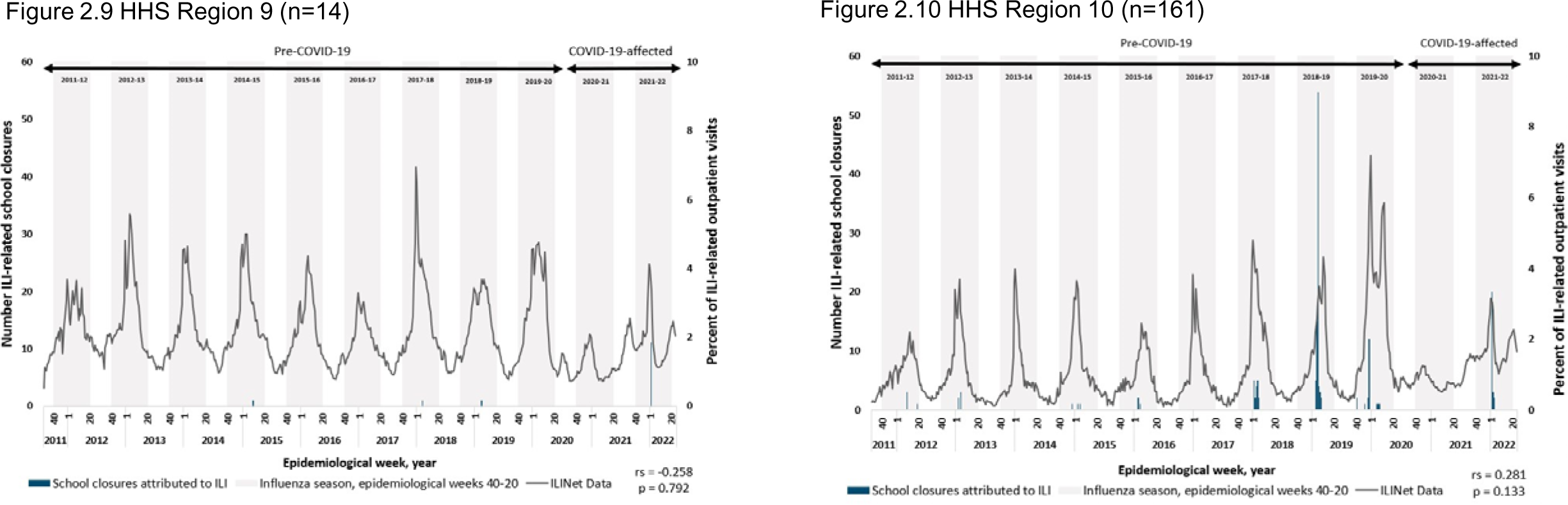
1-10. Estimated number of school closures associated with influenza-like illness (ILI) and percent of outpatient provider visits for ILI* by HHS region, by epidemiologic week – United States, August 1, 2011—June 30, 2022. * Percent of outpatient provider visits for ILI per data available from ILINet [6]. Note: In the 2012-13, 2014-15, 2016-17, and 2017-18 influenza seasons, influenza A (H3N2) was the predominant strain [8]. In the 2011-12 influenza season, which was unusually mild, influenza A (H3N2) predominated overall but, influenza A (H1N1)pdm09 and influenza B also widely circulated [8]. In the 2013-14 and 2015-16 seasons, influenza A (H1N1)pdm09 was the predominant strain [8]. In the 2018-19 season, there were two peaks of similar magnitude dominated by influenza A (H1N1) followed by influenza A (H3N2) [8]. During the 2019-2020 season, Influenza B predominated early in the season followed by influenza A (H1N1)pdm09 [8]. In the 2020-2021 season, there was unusual low flu activity in the United States when both Influenza A ((H1N1)pdm09)and (H3N2)) and influenza B, and the majority of influenza A viruses were H3N2 [8]. In the 2021-2022 season, the majority of positive flu tests reported to the CDC by US Public Health Laboratories were attributed to influenza A (H3N2) [8]

### Pre-COVID-19 school years (2011/12 to 2019/20)

Over the initial nine years of the study period, encompassing the 2011/12 to 2019/20 school years, 2,019 observed ILI-SC events accounted for an estimated total of 8,901 school closures (Table 1). The median duration of closure was 2 school days (range: 1–7) (Table S2). The largest number of ILI-SCs [2,886 (32·4%)] were captured in the 2019/20 school year prior to the onset of the COVID-19 pandemic in March 2020 (Table 1). The prior two school years, 2017/18 and 2018/19, also had high numbers with 1,966 (22·1%) and 1,912 (21·5%) ILI-SCs, respectively.

The timing of ILI-SC occurrence on the national level in relation to outpatient ILI activity varied across influenza seasons (Fig 1). The time series begins with the 2011/12 season, the mildest influenza season observed, which only reached the national baseline of percent of outpatient provider visits for ILI, as reported by ILINet, for a period of one week and all closures occurred after winter break (Fig S1). In the following three seasons, ILI activity breached the national baseline in late fall and peaked near the first of the year, while the peaks and majority of ILI-SCs followed a few weeks after. In the subsequent four seasons, relatively later peaks (in February and March) of ILI activity were preceded or met by the majority of ILI-SCs. During the 2019/20 season, ILI activity surpassed the national baseline in early November and was characterized by three peaks; the second of these, in early February, coincided with the peak of ILI-SCs. As school closed preemptively and almost synchronously nationwide as a countermeasure to the start of the COVID-19 pandemic [10], ILI activity sharply declined from early March; no additional ILI-related school closures were observed beyond March 11, 2020 (Fig 1, Fig S1).

The strongest correlations (> r_s_=0·80) between ILI-SCs and national outpatient healthcare visits for ILI occurred in the three seasons between 2016/17 and 2018/19, which were dominated, all or in-part, by influenza A (H3N2) (Table S3). Strong correlation was seen in the 2019/20 season, dominated in turn by both influenza A (H1N1) and influenza B, when truncated to the last observed ILI-SC in epidemiological week 11 (week of March 8, 2020) [r_s_=0·77 (p<0·001)]. Truncation at the preceding and following weeks led to lower correlations.

A moderate correlation was noted between the ILI-SCs and all-age laboratory-confirmed influenza-associated hospitalizations in the 13 states that routinely report these data [r_s_=0·56 (p<0·001); Fig 3, Table S4]. Correlation was slightly higher for those in the 5-17 years age group, which aligns with K-12 students, [r_s_=0·59 (p<0·001)] [Table S4].

**Fig 3.**
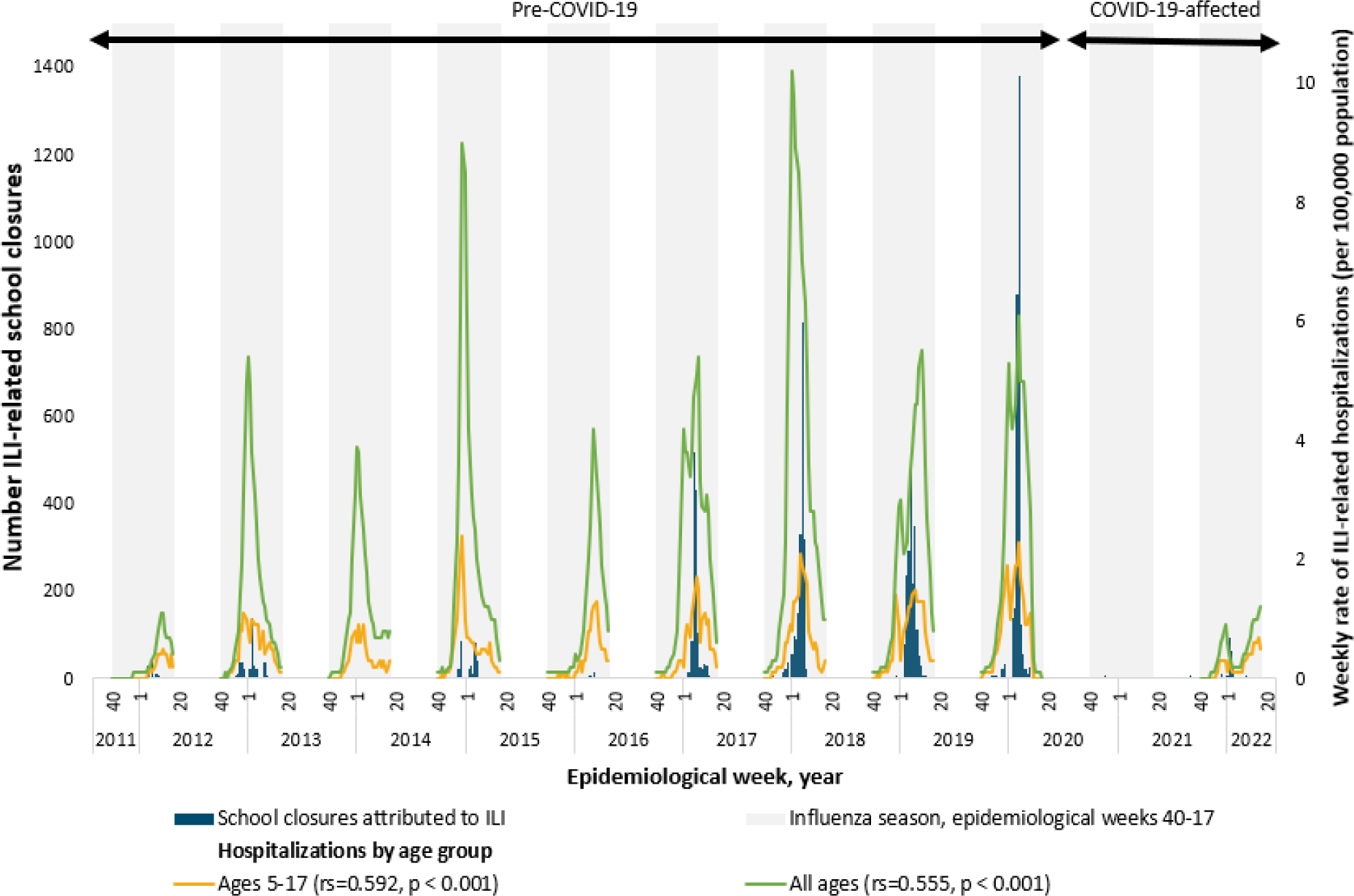
Estimated number of school closures associated with influenza-like illness and laboratory-confirmed influenza-associated hospitalizations* – 13 states^†^, August 1, 2011—June 30, 2022. *Data on laboratory-confirmed influenza-associated hospitalizations per data from FluSurv-NET [7] are shown for two groups, “all ages” and “ages 5-17”. Ages 5-17 reflect school-age children, which had the strongest correlation with influenza/ILI-related school closures. Lines for hospitalization data are not continuous because data are only reported for epidemiological weeks 40-17. ^†^Data on laboratory-confirmed influenza-associated hospitalizations [7] were available from the following states: CA, CO, CT, GA, MD, MN, NM, NY, OR, TN, MI, OH, and UT. Note: In the 2012-13, 2014-15, 2016-17, and 2017-18 influenza seasons, influenza A (H3N2) was the predominant strain [8]. In the 2011-12 influenza season, which was unusually mild, influenza A (H3N2) predominated overall but, influenza A (H1N1)pdm09 and influenza B also widely circulated [8]. In the 2013-14 and 2015-16 seasons, influenza A (H1N1)pdm09 was the predominant strain [8]. In the 2018-19 season, there were two peaks of similar magnitude dominated by influenza A (H1N1) followed by influenza A (H3N2) [8]. During the 2019-2020 season, Influenza B predominated early in the season followed by influenza A (H1N1)pdm09 [8]. In the 2020-2021 season, there was unusual low flu activity in the United States when both Influenza A ((H1N1)pdm09)and (H3N2)) and influenza B, and the majority of influenza A viruses were H3N2 [8]. In the 2021-2022 season, the majority of positive flu tests reported to the CDC by US Public Health Laboratories were attributed to influenza A (H3N2) [8] Note: Spearman rank correlations were used to evaluate the relationship between Influenza/ILI-related school closures and laboratory-confirmed influenza-associated hospitalizations during influenza seasons (epidemiological weeks 40 through 17).

### COVID-19-affected school years (2020**/**21 to 2021**/**22)

In the two complete school years following the onset of the COVID-19 pandemic, 2020/21 and 2021/22, 235 schools closed as part of 58 ILI-SC events (Table 1). All ILI-SCs in 2020/21 (n=11) and 200 (89·3%) in 2021/22 were attributed to both ILI and COVID-19 (Table S5). ILI-SCs in the 2020/21 school year were the longest in the study period, with a median of 4 days (range 3–33) as compared to medians of 1–2 days in all other years including 2021/22 (Table S2).

In the 2020/21 season, ILI activity did not surpass the national baseline and all ILI-SCs preceded the winter holidays (Fig S1). In the 2021/22 season, ILI activity breached the national baseline but for fewer weeks as compared to all pre-COVID-19 seasons except 2011/12, and ILI-SCs reached their height a couple weeks after the ILI peak. The weakest correlation between annual ILI-SCs and national outpatient healthcare visits was observed in 2020/21[0·29 (p=0·103)] (Table S3). Likewise, correlation between ILI-SCs and laboratory-confirmed influenza-associated hospitalizations were weak for all ages [0·32 (p=0·011)] and not significant for the 5-17 age group during the COVID-19-affected period [0·22 (p=0·087)] (Table S4).

## Discussion

Our data demonstrate that ILI-SCs occur annually in the US with temporality that reflects the general patterns of influenza activity on both national and regional levels as observed through routine surveillance of medically attended ILI. In most seasons prior to the COVID-19 pandemic, school winter breaks temporarily reduced ILI activity with seasonal peaks usually following within a few weeks of the start of second semester. ILI-SC annual peaks occurred in the same timeframe, presumably once the number of ill students reached a critical mass a few weeks after school re-opening for spring semester. A similar pattern was exhibited around the winter break of the 2019/20 school year; however, the advent of the COVID-19 pandemic and subsequent preemptive closure of nearly all K-12 schools in the US likely impacted the remaining weeks of both the season’s ILI activity and related SCs. During the subsequent two COVID-affected years, 2020/21 and 2021/22, ILI-SCs reflected the temporality and low levels of ILI activity.

Prior to the COVID-19 pandemic, the highest numbers of ILI-SCs occurred in seasons when influenza A (H3N2) was predominant either nationally or regionally. Among these, the 2017/18 and 2018/19 seasons each reached a level of closures previously only seen in the 2009 pandemic where a reported 1,947 schools closed [11]. Meanwhile, exceptionally mild seasons, such as 2011/12 or those dominated by influenza A (H1N1) generally had far fewer closures.

However, the largest annual number of ILI-SCs during the study period occurred during the 2019/20 school year, which was affected by an intense seasonal outbreak predominated first by influenza B and subsequently by influenza A (H1N1). This record was set despite the school year being abruptly interrupted, by the implementation of measures used to mitigate the spread of SARS-CoV-2 as early as February 2020 and ultimately by the near simultaneous preemptive closure of all K-12 schools beginning in the week ending March 14, 2020 in order to slow transmission of SARS-CoV-2 [10,12].

In addition to variations in seasonal influenza epidemiology, the number of ILI-SCs per year may have been influenced by differences in seasonal vaccine effectiveness. The higher counts of ILI-SCs in H3N2-dominated years may, at least in part, be related to a relatively higher effectiveness of the influenza A (H1N1) vaccine component compared with the A (H3N2) component [13]. Furthermore, because the predominant circulating virus types and subtypes can vary geographically [6], vaccine effectiveness may also differ by locale.

Though vaccine effectiveness may be a contributing factor to seasonal distribution of ILI-SC occurrence, uneven geographic distribution could also have been related to variations in vaccine uptake or certain programmatic factors that prompt school authorities to close schools, which may be common to a state or locale.

There were observable regional differences in vaccine coverage across the study period. Among HHS Regions with relatively few ILI-SCs across the study period, particularly regions 1, 2, 3, 7, and 8, estimated vaccine coverage across the study period tended to be higher than national estimates. Meanwhile HHS Region 4, the region with the most ILI-SCs, reported vaccine coverage estimates consistently below the national-level [14]. This may suggest that seasonal vaccine uptake is a contributing factor in ILI-SC occurrence, and, while not a key driver, potentially has a bigger impact in some years than others.

There are likely also programmatic factors that impact school closure decisions. For example, regional variability could be affected by differences in state policies related to school funding or the varying costs to run schools. Student enrollment is typically a primary driver of funds provided to schools by the state, and, depending on how the number of enrolled students is calculated in each state, school closure decisions may be swayed by illness-related absenteeism given the connection between attendance and funding [15]. Additionally, factors such as the higher costs of food service and transportation in rural schools [16] may impact the cost-effectiveness of running differently located schools with high levels of illness-related absenteeism. More research is needed to analyze the effect of policies and programmatic factors on the reasons for and barriers to closure. Disproportionate burden of ILI-SCs appears to be concentrated in socio-economically disadvantaged communities of two HHS regions, which adds urgency to further evaluate and, where needed, address programmatic factors that may be influencing these trends.

During the COVID-dominated period (school years 2020/21 and 2021/22), ILI-SC data collection continued but remarkably few ILI-SCs were documented. The 2020/21 school year had among the lowest number of ILI-SCs, comparable only to the 2013/14 school year, a moderate influenza A (H1N1)-dominated season. Moreover, all documented ILI-SCs in the 2020/21 school year were simultaneously attributed to COVID-19, and the weight of each disease’s contribution to the closure decisions is unknown. The low number of observed ILI-SCs in the 2020/21 school year is credible, because the ILI activity that year did not breach the national baseline, a first during the study period.

While the 2021/22 school year saw an increase in ILI-SCs from the previous year, the numbers were only a fraction of the number of ILI-SCs in the three school years prior to the COVID-19 pandemic, all dominated by influenza A (H3N2). This pattern is consistent with the generally low ILI activity observed during the COVID-dominated years [12].

In response to the growing levels of community SARS-CoV-2 transmission in the summer and fall of 2020, many schools and districts across the country offered a variety of reduced-contact learning options in the 2020/21 school year including hybrid (partly in-person / partly distance learning) and fully virtual (online) education models, thereby limiting the density, frequency, and duration of in-person student congregation in schools [17]. In schools where in-person learning did take place, various measures such as social distancing, facemasks, and increased cleaning were recommended to reduce transmission of SARS-CoV-2 [18]. The nearly total return to in-person learning in the 2021/22 school year [19] coincided with an uptick in both ILI-activity and ILI-SCs.

ILI-SCs described in this multi-year study are reactive closures associated with increased ILI activity, i.e. they occurred as a consequence of increased disease transmission as reported in most school closure announcements. In contrast, during the 2009 Influenza A (H1N1) pandemic, both preemptive and reactive closures were implemented in the US and other countries [20]. Subsequently performed systematic literature reviews support pre-emptive school closures as an NPI to reduce virus spread in schools and surrounding communities [21, 22], but the effectiveness of closures implemented reactively remains unclear. Two US-based studies reported that reactive closures have little effect during seasonal [23] or pandemic [24] influenza outbreaks; conversely, studies implemented elsewhere indicate that transmission would be even greater if they are not implemented [25, 26], though some with only minor effect [26]. Reactive closures have for decades been routinely used in Japan and Russia to control outbreaks of seasonal and pandemic influenza where they are triggered locally by defined levels of heightened ILI-related student absenteeism [25, 27]. However, it has yet to be shown under what conditions reactive closures might help control local outbreaks in the US. Meanwhile, recent US-based research has shown that routine monitoring of cause-specific student absenteeism can act as an early warning signal of increased ILI activity in schools and surrounding community [28]. For communities which are disproportionally burdened year-after-year with ILI-SCs, it may be particularly worthwhile to explore more effective alternatives to late reactive ILI-SCs, even if only via better (earlier) timing of the interruptions of in-person instruction to obtain greater reduction of within-school disease transmission.

This evaluation is subject to at least four limitations. First, ILI-SC data only captured closures reported through publicly available online media sources, potentially missing announcements made exclusively through other communication means (e.g. text messages, listservs, etc.). Second, because data points for ILI-SCs were abstracted exclusively from publicly available announcements, some information may not have been complete or entirely accurate. However, the method of data collection imposed no reporting burden on schools and was relatively inexpensive. Third, data on outpatient healthcare provider visits for ILI and laboratory-confirmed influenza hospitalizations were subject to limitations that are characteristic of surveillance systems. Additionally, not all states were included in hospitalization data and therefore results are not generalizable. However, this was mitigated by the fact that Tennessee, the state most greatly impacted by ILI-SCS, was accounted for in the hospitalization data. Lastly, we compared the relationships of ILI-SCs and ILI surveillance data on national and sub-national levels. Such high level of comparisons may not have fully reflected relationships on the local level (i.e., communities around closed schools), and further studies exploring these relationships on the local level are warranted.

To our knowledge, this study is the first of its kind in the US, describing the frequency and characteristics of school closures attributed to seasonal influenza/ILI over eleven consecutive influenza seasons, and thereby establishing an inter-influenza-pandemic baseline of such closures. This data collection also provided real-time situational awareness information during severe seasonal influenza outbreaks and, after suitable modification, was instrumental in documenting COVID-19-associated preemptive and reactive K-12 school closures in the United States from February 2020 through June 2022 [10]. The ILI-SC data, systematically collected from public sources, have been congruent with trends observed via national disease surveillance systems both during high ILI activity seasons as well as during the unusually low ILI incidence in COVID-affected years. Of note, during the study implementation ILI-SC data were available about a week sooner than ILINet data, i.e. ILI-SC data were being compiled in near-real time.

## Conclusion

Given that ILI-SC announcements are publicly available and, as demonstrated in course of this research, amenable to compiling in near-real time, the routine monitoring of ILI-SCs may be a useful addition to existing influenza surveillance systems, particularly during severe influenza seasons or pandemics when ILI-SCs are likely to occur more frequently. An alternative approach would be to initiate direct data collection from schools via electronic reporting of disease-related or indeed all unplanned school closures lasting >=1 day to state education agencies and organize data sharing at the federal level, but that would likely require a significantly greater effort. The reported economic burden of ILI-SCs documented by this study in the pre-COVID period is disproportionally concentrated in socio-economically disadvantaged parts of two HHS regions [29], which further underscores the importance of continued ILI-SC monitoring and research as we prioritize addressing health disparities. Meanwhile, given that ILI-SCs appear to be a drastic measure with little public health benefit for affected communities because they are undertaken by school authorities reactively, i.e. when within-school ILI activity has already reached critical proportions resulting in high student absenteeism, ways to reduce ILI disease burden in schools merit continued exploration. A multi-pronged approach that includes increased student and staff vaccination, better ventilation, and timely implementation of NPIs in combination with the monitoring of cause-specific absenteeism, could provide opportunities to timely reduce ILI transmission in schools before late reactive school closure becomes necessary.

## Data Availability

All data produced in the present study will be made available upon acceptance by a peer review journal.

## Disclaimer

The opinions expressed by authors contributing to this journal do not necessarily reflect the opinions of the Centers for Disease Control and Prevention or the institutions with which the authors are affiliated.

## Acknowledgments

The authors would like to thank the various data collection teams over the study period, including those with Oak Ridge Associated Universities (Oak Ridge, TN), the Mayatech Corporation (Silver Spring, MD), and our unit ORISE Fellows (Ashley Jackson, Cassandra Kersten, Jeffrey Hodis, Peter Kim, Sarah Moreland, Livvy Shafer, and Pallavi Malla) and contractors (Tamara Cummings, Jasminn Evans, Atea Francis, Elisha Gilbert, and Zaneta Oliver). For contributions to data visualization, we again thank Sarah Moreland, and for contributions to database management and quality control, we thank Jianrong Shi.

