## Supplementary Material for "School Closures due to Seasonal Influenza: Experience from Eleven Influenza Seasons – United States, 2011–2022"

**Authors**

Nicole Zviedrite^1*^, Ferdous Jahan^1, 2^, Yenlik Zheteyeva^1,3^, Hongjiang Gao^1^, Amra Uzicanin^1^

**Affiliations**

^1^Centers for Disease Control and Prevention, Atlanta, Georgia, USA.

^2^Cherokee Nation Operational Solutions, LLC, Tulsa, Oklahoma, USA

^3^CDC/DGMQ until January 2018. Present affiliation: : Daiichi Sankyo, Inc.

**Corresponding author***

Nicole Zviedrite, MPH, Community Interventions for Infection Control Unit, Division of Global Migration and Quarantine, National Center for Emerging Zoonotic Infectious Diseases, Centers for Disease Control and Prevention, 1600 Clifton Rd NE, MS V18-2, Atlanta, GA 30329,, 404-639-3961

**[Table S5. Publicly announced, reactive* school closures associated with illness,](#_Toc129360834) [by school year – United States, August 1, 2011—June 30, 2022](#_Toc129360834)**	[18](#_Toc129360834)

**[Figure S2. Estimated number of school closures associated with illness](#_Toc129360835)^[*](#_Toc129360835)^ [(N=58,015) and percent of outpatient provider visits for ILI](#_Toc129360835)^[†](#_Toc129360835)^ [by epidemiologic week – United States, August 1, 2011—June 30, 2022](#_Toc129360835)**	[19](#_Toc129360835)

**[Table S6. Selected characteristics of influenza-like illness-related school closures — United States, August 1, 2011—June 30, 2022](#_Toc129360837)**	[21](#_Toc129360837)

**[Table S7. Recurrence of influenza-like illness-related school closures within each school year among unique schools](#_Toc129360839)^[*](#_Toc129360839)^[, United States, August 1, 2011 – June 30, 2022](#_Toc129360839)**	[23](#_Toc129360839)

**[Table S8. Characteristics of schools with repeat closures due to influenza-like illness by school year -- United States, August 1, 2011—June 30, 2022](#_Toc129360840)**	[24](#_Toc129360840)

**[Table S9. Recurrence of influenza-like illness (ILI)-related school closures among unique schools](#_Toc129360841)^[*](#_Toc129360841)^[– United States, August 1, 2011 – June 30, 2022](#_Toc129360841)**	[26](#_Toc129360841)

### **ADDITIONAL DESCRIPTION OF METHODS**

To supplement the methods section, additional information is provided below. Included are methods to describe reasons for school closure, to describe the characteristics of schools closed for ILI, to conduct bivariate and multiple logistic regression analysis on school characteristics, as well as to conduct descriptive analyses of repeat closures for ILI and ILI closures in the context of all disease-related closures.

#### CATEGORIZATION OF REASONS FOR CLOSURE

##### **Table S1.** **Approach to grouping quotes from original school closure announcements to define influenza-/illness-related closures and two additional categories of factors related to illness**

| **Example Direct Quotes as Published in the School Closure Announcement** | **Categories** |
| --- | --- |
| **Main Categories** |  |
| - “Due to flu” - “Due to influenza” - “Due to flu-like illness” | Announcement mentions only flu/illness |
| **Additional Categories** |  |
| **Student and Staff Absenteeism** |  |
| - “There are so many sick teachers and students that they decided to shut down school today.” - “Due to increased illness among many of our teachers and students, [school] will be closed on Monday…” | Due to increased number of ill students/staff |
| - “School will not be in session tomorrow, [] due to the large amount of illness related student absences.” - “In a Facebook post [] School in [] says they have closed the school due to a very high rate of illness and absence among both students and faculty/staff.” | Due to increased student absenteeism |
| - “Many faculty members are sick and officials could not find enough substitutes.” - “School officials don’t believe they will have enough staff members to open school, and while they have a number of students ill as well, it appears to be more staff members than students.” - “We did not have enough substitutes for today and its looks worse for tomorrow.” | Due to lack of teachers/substitutes due to illness |
| **Intervention or programmatic factors** |  |
| - “The closure is an attempt to stop the spread of influenza-related illness that has run rampant through the school.” - “She felt she had to cancel school to prevent infecting the rest of the students, she said.” - “The [school district] is closing Friday, as a precautionary measure, after a student tested positive for H1N1 strain of the flu.” | To stop/halt/prevent illness spread |
| - “Due to flu, 2 pm dismissal today and classes cancelled canceled canceled Tuesday & Wednesday for deep cleaning.” - “Students at … Elementary School … will not be attending class until the building is disinfected to prevent further outbreaks, the district said.” | To clean/disinfect classrooms, buildings, and facilities |
| - “… superintendent [] decided to cancel classes for Tuesday and Wednesday so dozens of students could recover from the flu-like symptoms.” - “The school will shut down Thursday and Friday to give students and teachers time to rest and allow janitors to clean the school.” | To allow students/teachers to rest and recover |
| - “State funding for school districts is based on daily attendance, meaning that continuing to provide full instruction days when attendance is low will reduce the amount of funding received from the state, [they] said… [They] decided to cancel school for state funding purposes and to reduce the spread of the flu, strep throat and other illnesses the district has been seeing lately, he said.” - “This isn't just a health issue. It's also an economic one. Every day, the school gets $28.50 per student that attends class. Today, 61 students were out, costing the school more than $1,700. The small district says they can't afford to lose anymore.” - “Officials said funding from the state is based on average daily attendance and that keeping the schools open would have cost the district money.” | Due to financial reasons |

Note: These are direct quotes from ILI-SC announcements identified by this project. Identifying information of schools and school districts have been replaced by empty brackets (“[ ]”)

#### DESCRIPTIVE CHARACTERISTICS OF ILI-SCS

In addition to NCES variables discussed in the methods section, we also downloaded data related to other school characteristics, including the number of students enrolled in the federal free or reduced-price school lunch program, urbanicity, and student-teacher ratio (1, 2). We produced descriptive statistics using both the variables mentioned in the main methods and those mentioned here to describe the characteristics of schools which closed for ILI.

#### ANNUAL TIMELINES

We disaggregated the full study period timeline to show annual (by school year/influenza season) estimated counts of ILI-SCs with corresponding reported percent of outpatient provider visits for ILI and the annual baseline as reported by ILINET (3). ILI-SCs for HHS Region are shown separately from all other HHS Regions (4).

#### BIVARIATE AND MULTIPLE LOGISTIC REGRESSION

Using bivariate and multiple logistic regression, we examined the associations between ILI-SCs and certain school characteristics in the NCES data set (urbanicity, student-teacher ratio, and percentages of students eligible for subsidized school meals) for public schools (1), which comprise 96.4% of schools in our data set. Private schools accounted for the remaining 3.6% of ILI-SCs, and were excluded as the NCES Private School Survey data (2) did not fully include the corresponding variables found in public school data.

#### REPEAT CLOSURES FOR ILI

During the course of analysis, repeat closures of the same schools for ILI were noted in the data, occurring both within and across school years. We describe the patterns seen across the study period.

#### ALL DISEASE CLOSURES

To better understand ILI-SCs in the broader context of other types of reactive, disease-related closures which occurred during the period, we examine a broader subset of data. These data include all reactive school closure for which any illness or disease was reported as a reason for the closure decision.

### **SUPPLEMENTARY RESULTS**

#### CHARACTERISTICS OF ILI-SCS

The overwhelming majority of ILI-SCs in the pre-COVID-19 period (95·5%) occurred consistently during winter months (December-February) (Table S2). Elementary and elementary-middle schools had the largest number of closures for pre-COVID years, with 3,080 (34·6%) and 2,103 (23·6%), respectively. During pre-COVID years ILI-SCs were more frequently in rural areas [4,700 (52·8%)] as compared to town [1,998 (22·4%)], urban [1,225 (13·8%)], and suburban [956 (10·7%)] areas.

See Table S2.

##### **Table S2.** **Characteristics of school closures associated with influenza-like illness by school year -- United States, August 1, 2011—June 30, 2022**

|  | **School year** | | | | | | | | | | | | | |
| --- | --- | --- | --- | --- | --- | --- | --- | --- | --- | --- | --- | --- | --- | --- |
|  | **Pre-COVID-19 Years^*^**^#^ | | | | | | | | | | **COVID-19-affected Years**^#^ | | | **Total**^#^ |
|  | **2011-12** | **2012-13** | **2013-14** | **2014-15** | **2015-16** | **2016-17** | **2017-18** | **2018-19** | **2019-20** | **Subtotal** | **2020-21** | **2021-22** | **Subtotal** |  |
| Estimated Number of schools closed for influenza/ILI^†^, n (%) | 103 (1) | 383 (4) | 11 (0) | 308 (3) | 40 (0) | 1292 (14) | 1,966 (22) | 1,912 (21) | 2,886 (32) | 8,901 | 11 (0) | 224 (2) | 235 | 9,136 |
| Season, n (%) |  |  |  |  |  |  |  |  |  |  |  |  |  |  |
| Fall (Sep-Nov) | 0 | 26 (7) | 0 | 7 (2) | 0 | 0 | 8 (0) | 2 (0) | 30 (1) | 73 (1) | 11 (100) | 16 (7) | 27 (11) | 100 (1) |
| Winter (Dec-Feb) | 86 (84) | 323 (84) | 11 (100) | 298 (97) | 19 (48) | 1,194 (92) | 1,956 (99) | 1,794 (94) | 2,818 (98) | 8,499 (95) | 0 | 196 (88) | 196 (83) | 8,695 (95) |
| Spring (Mar-May) | 17 (17) | 34 (9) | 0 | 3 (1) | 21 (53) | 98 (8) | 2 (0) | 116 (6) | 38 (1) | 329 (4) | 0 | 12 (5) | 12 (5) | 341 (4) |
| Summer (Jun-Aug) | 0 | 0 | 0 | 0 | 0 | 0 | 0 | 0 | 0 | 0 | 0 | 0 | 0 | 0 |
| School type, n (%) |  |  |  |  |  |  |  |  |  |  |  |  |  |  |
| Public Schools | 92 (89) | 361 (94) | 9 (82) | 279 (91) | 33 (83) | 1,255 (97) | 1,887 (96) | 1,852 (97) | 2,803 (97) | 8,571 (96) | 11 (100) | 223 (100) | 234 (100) | 8,805 (96) |
| Private schools | 11 (11) | 22 (6) | 2 (18) | 29 (9) | 7 (18) | 37 (3) | 79 (4) | 60 (3) | 83 (3) | 330 (4) | 0 | 1 (0) | 1 (0) | 331 (4) |
| School level, n (%) |  |  |  |  |  |  |  |  |  |  |  |  |  |  |
| Elementary school (K-5 grade) | 17 (17) | 95 (25) | 5 (45) | 78 (25) | 6 (15) | 466 (36) | 741 (38) | 674 (35) | 998 (35) | 3,080 (35) | 5 (45) | 93 (42) | 98 (42) | 3,178 (35) |
| Elementary-middle school (K-8 grade) | 49 (48) | 135 (35) | 2 (18) | 103 (33) | 12 (30) | 298 (23) | 386 (20) | 413 (22) | 705 (24) | 2,103 (24) | 1 (9) | 34 (15) | 35 (15) | 2,138 (23) |
| Elementary-high school (K-12 grade) | 6 (6) | 14 (4) | 0 | 16 (5) | 8 (20) | 76 (6) | 112 (6) | 110 (6) | 157 (5) | 499 (6) | 0 | 4 (2) | 4 (2) | 503 (6) |
| Middle school (6-8 grade) | 7 (7) | 39 (10) | 2 (18) | 35 (11) | 4 (10) | 151 (12) | 239 (12) | 223 (12) | 322 (11) | 1,022 (11) | 2 (18.2) | 31 (14) | 33 (14) | 1,055 (12) |
| Middle-high school (6-12 grade) | 11 (11) | 30 (8) | 1 (9) | 21(7) | 4 (10) | 87 (7) | 126 (6) | 163 (9) | 214 (7) | 657 (7) | 0 | 17 (8) | 17 (7) | 674 (7) |
| High school (9-12 grade) | 13 (13) | 69 (18) | 1 (9) | 55 (18) | 6 (15) | 213 (16) | 361 (18) | 322 (17) | 477 (17) | 1,517 (17) | 3 (27) | 42 (19) | 45 (19) | 1,562 (17) |
| Type of school not specified | 0 | 1 (0) | 0 | 0 | 0 | 1 (0) | 1 (0) | 7 (0) | 13 (0) | 23 (0) | 0 | 3 (1) | 3 (1) | 26 (0) |
| School setting, n (%) |  |  |  |  |  |  |  |  |  |  |  |  |  |  |
| Rural | 72 (70) | 301 (79) | 10 (91) | 199 (65) | 19 (48) | 646 (50) | 941 (48) | 977 (51) | 1,535 (53) | 4,700 (53) | 1 (9) | 100 (45) | 101 (43) | 4,801 (53) |
| Town | 7 (7) | 59 (15) | 0 | 73 (24) | 11 (28) | 297 (23) | 440 (22) | 443 (23) | 668 (23) | 1,998 (22) | 9 (82) | 68 (30) | 77 (33) | 2,075 (23) |
| Suburb | 14 (14) | 10 (3) | 0 | 22 (7) | 7 (18) | 172 (13) | 238 (12) | 189 (10) | 304 (11) | 956 (11) | 1 (9) | 30 (13) | 31 (13) | 987 (11) |
| City | 10 (10) | 12 (3) | 1 (9) | 14 (5) | 3 (8) | 176 (14) | 346 (18) | 296 (15) | 367 (13) | 1,225 (14) | 0 | 25 (11) | 25 (11) | 1,250 (14) |
| Not specified | 0 (0) | 1 (0) | 0 | 0 (0) | 0 (0) | 1 (0) | 1 (0) | 7 (0) | 12 (0) | 22 (0) | 0 | 1 (0) | 1 (0) | 23 (0) |
| HHS Region^‡^, n (%) |  |  |  |  |  |  |  |  |  |  |  |  |  |  |
| Region 1 | 1 (1) | 0 | 0 | 0 | 1 (3) | 1 (0) | 5 (0) | 1 (0) | 5 (0) | 14 (0) | 0 | 7 (3) | 7 (3) | 21 (0) |
| Region 2 | 0 | 0 | 3 (27) | 2 (1) | 1 (3) | 3 (0) | 7 (0) | 4 (0) | 23 (1) | 43 (0) | 0 | 4 (2) | 4 (2) | 47 (1) |
| Region 3 | 0 | 7 (2) | 0 | 6 (2) | 0 | 19 (1) | 4 (0) | 38 (2) | 115 (4) | 189 (2) | 0 | 0 (0) | 0 (0) | 189 (2) |
| Region 4 | 64 (62) | 274 (72) | 1 (9) | 212 (69) | 20 (50) | 1,061 (82) | 1,155 (59) | 1,345 (70) | 1,882 (65) | 6,014 (68) | 1 (9) | 25 (11) | 26 (11) | 6,040 (66) |
| Region 5 | 9 (9) | 38 (10) | 7 (64) | 29 (9) | 13 (33) | 114 (9) | 245 (12) | 38 (2) | 449 (16) | 942 (11) | 1 (9) | 48 (21) | 49 (21) | 991 (11) |
| Region 6 | 0 | 46 (12) | 0 | 46 (15) | 1 (3) | 60 (5) | 441 (22) | 339 (18) | 364 (13) | 1,297 (15) | 9 (82) | 77(34) | 86 (37) | 1383(15) |
| Region 7 | 22 (21) | 11 (3) | 0 | 6 (2) | 1 (3) | 25 (2) | 86 (4) | 54 (3) | 20 (1) | 225 (3) | 0 | 27 (12) | 27 (11) | 252 (3) |
| Region 8 | 3 (3) | 2 (1) | 0 | 3 (1) | 0 | 9 (1) | 4 (0) | 9 (0) | 8 (0) | 38 (0) | 0 | 0 (0) | 0 (0) | 38 (0) |
| Region 9 | 0 | 0 | 0 | 1 (0) | 0 | 0 | 1 (0) | 1 (0) | 0 | 3 (0) | 0 | 11 (5) | 11 (5) | 14 (0) |
| Region 10 | 4 (4) | 5 (1) | 0 | 3 (1) | 3 (8) | 0 | 18 (1) | 83 (4) | 20 (1) | 136 (2) | 0 | 25 (11) | 25 (11) | 161 (2) |
| Number of closure days ^§^, median (range) | 1 (1-2) | 1 (1-4) | 1 (1-1) | 1 (1-3) | 1 (1-3) | 2 (1-6) | 2 (1-7) | 2 (1-6) | 2 (1-6) | 2 (1-7) | 4 (3-33) | 2 (1-9) | 2 (1-33) | 2 (1-33) |
| Closures that lasted ≥4 days, n (%) | 0 | 21 (5) | 0 | 0 | 0 | 155 (12) | 105 (5) | 155 (8) | 150 (5) | 586 (7) | 7 (64) | 21 (9) | 28 (12) | 614 (7) |
| Percent students eligible for free/reduced-price lunch, median (IQR) ^¶^ | 74 (60-83) | 68 (59-77) | 57 (15-60) | 68 (58-79) | 63 (49-70) | 66 (55-79) | 63 (50-75) | 63 (50-75) | 59 (47-71) | 63 (50-74) | 69 (60-76) | 53 (40-69) | 53 (41-69) | 62 (50-74) |

^*^The 2019-20 school year is included in pre-COVID-19 seasons and include ILI-SCs through epidemiological week 11 of 2020, the last week of an ILI-related closure in the school year. While first COVID-19-related school closure occurred during epidemiological week 9 in 2020, widespread COVID-19-related schools closures were reported during the subsequent epidemiological weeks 12 and 13 of 2020 (5).

^†^Schools were counted once for each time they were part of a school closure event at either the district-level or school level. Number of schools in district-level closure events were estimated based on the number of K-12 schools in each affected school district per data available from the National Center for Education Statistics (1).

^‡^Regions of the United States Department of Health & Human Services (HHS) (4).

^§^759 closures did not have reopening dates and were defaulted to 1 day closure.

^¶^Private schools were excluded from this part of the analysis (2), the data is applicable for public schools only (1).

^#^Percentages may not add up to 100%, as they are rounded to the nearest percent.

#### ANNUAL TIMELINES OF ILI-SCS

##### **Figure S1. Comparison of estimated number of school closures associated with influenza-like illness in HHS region 4 (blue) vs. other HHS regions (green), per season, by epidemiologic week – United States, August 1, 2011—June 30, 2022^*^**

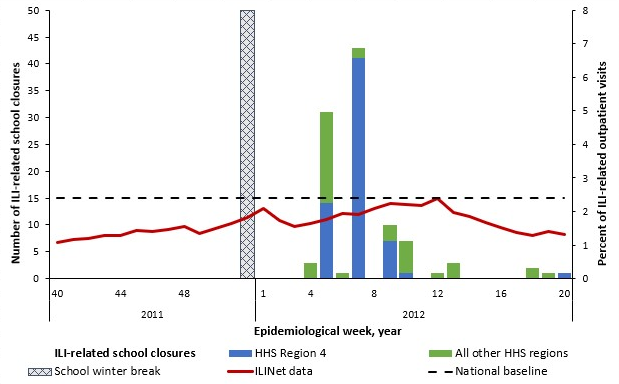

Figure S1.1 2011 – 2012 Influenza Season (n=103)

Season dominated by influenza A (H3N2)^†^

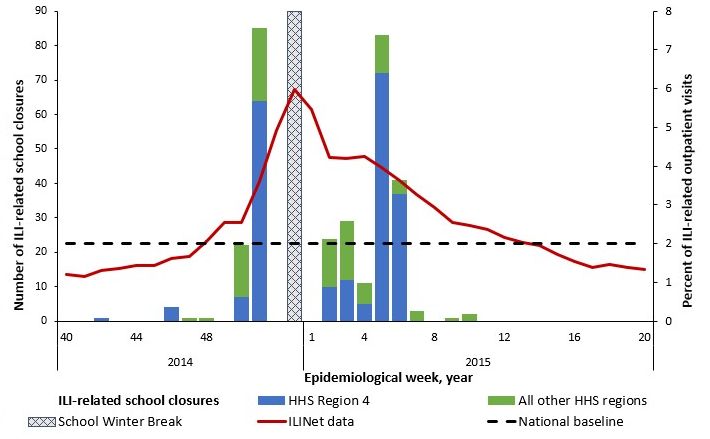

Figure S1.4 2014 – 2015 Influenza Season (n=308)

Season dominated by influenza A (H3N2)^†^

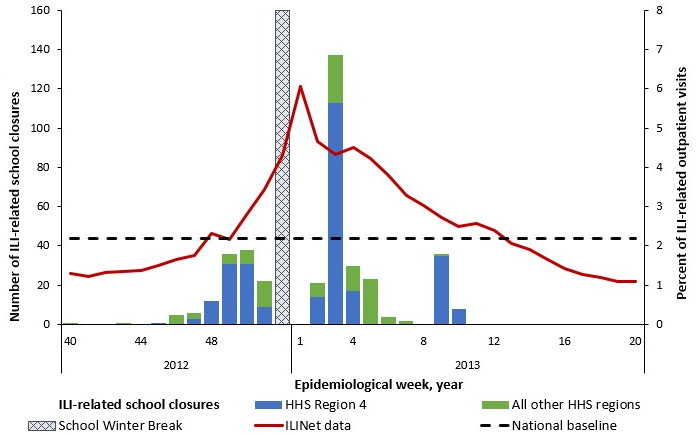

Figure S1.2 2012 – 2013 Influenza Season (n=383)

Season dominated by influenza A (H3N2)^†^

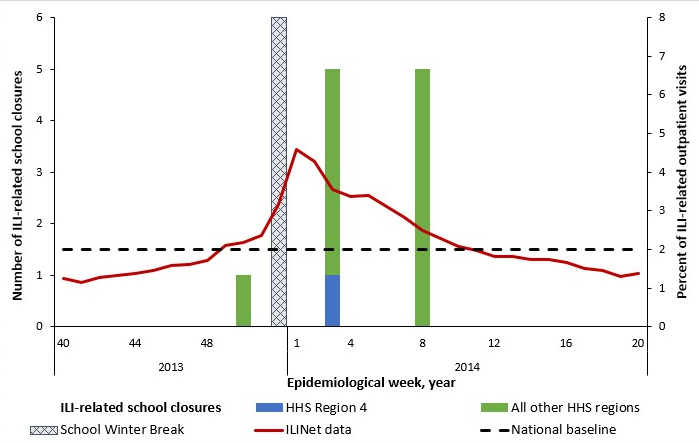

Figure S1.3 2013 – 2014 Influenza Season (n=11)

Season dominated by influenza A (H1N1)pd09^†^

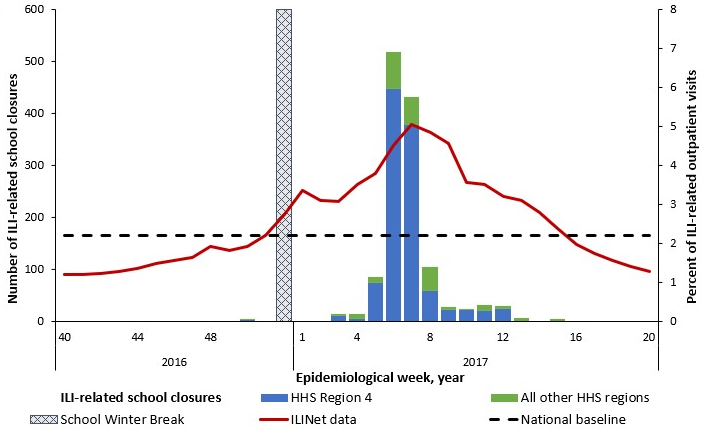

Figure S1.6 2016 – 2017 Influenza Season (n=1,292)

Season dominated by influenza A (H3N2)^†^

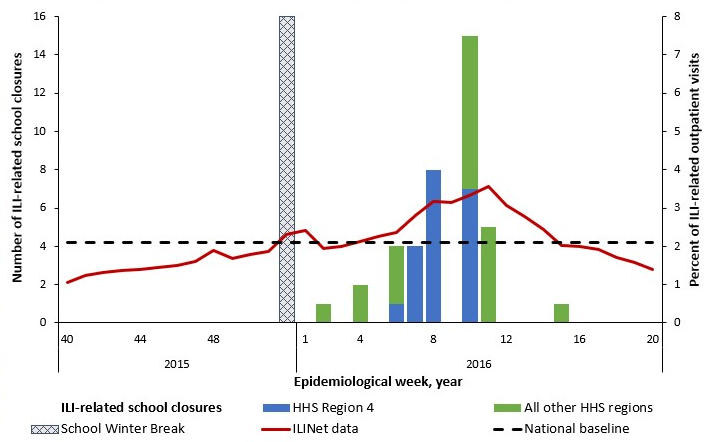

Figure S1.5 2015 – 2016 Influenza Season (n=40)

Season dominated by influenza A (H1N1)pd09^†^

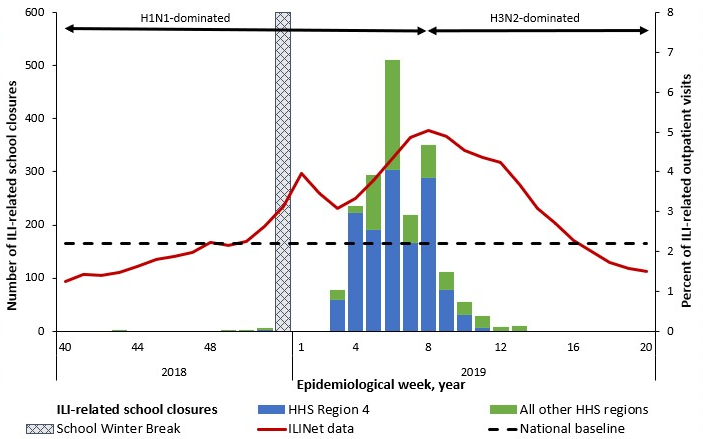

Figure S1.8 2018 – 2019 Influenza Season (n=1,912)

Season dominated by influenza A (H1N1) and influenza A (H3N2)^†^

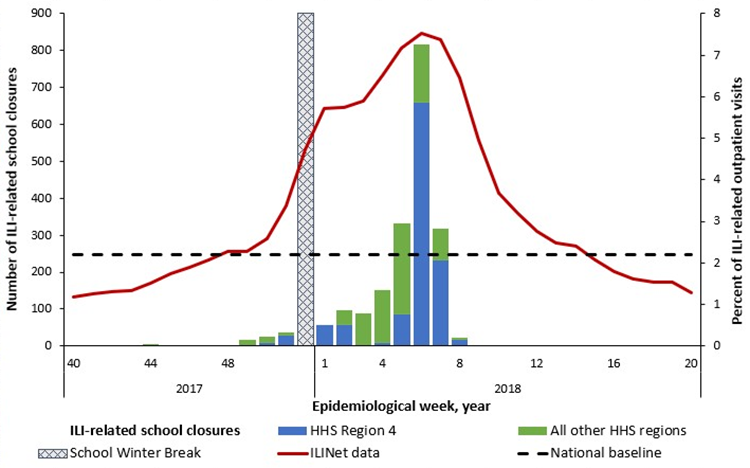

Figure S1.7 2017 – 2018 Influenza Season (n=1,966)

Season dominated by influenza A (H3N2)^†^

Figure S1.10 2020 – 2021 Influenza Season (n=11)

Season dominated by influenza A (H3N2)^†^

Season dominated by influenza B and influenza A (H1N1)^†‡^

Figure S1.9 2019 – 2020 Influenza Season (n=2,886)

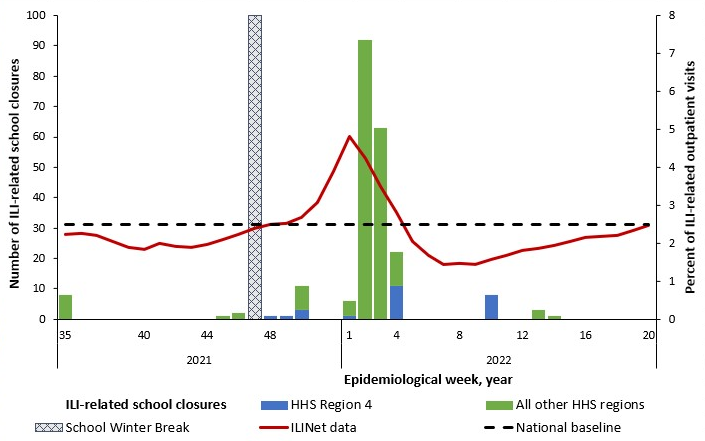

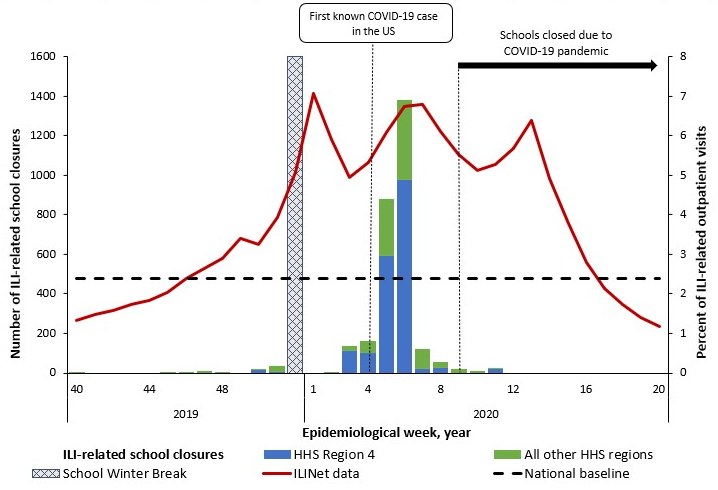

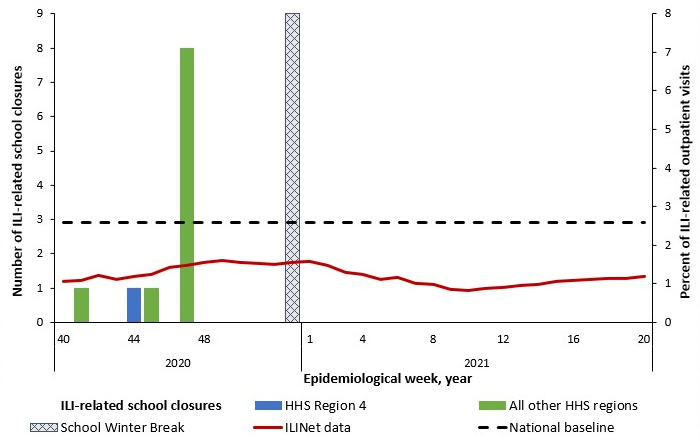

Figure S1.11 2021 – 2022 Influenza Season (n=224)

Season dominated by influenza A (H3N2)^†^

^*For context, the national percentage of outpatient provider visits for ILI per data available from ILI Net (3) is being shown as a solid red line and the national baseline is shown as a dotted black line and measured along the secondary Y-axis.^

^†^ ^In the 2012-13, 2014-15, 2016-17, and 2017-18 influenza seasons, influenza A (H3N2) was the predominant strain (6). In the 2011-12 influenza season, which was unusually mild, influenza A (H3N2) predominated overall but, influenza A (H1N1)pdm09 and influenza B also widely circulated. In the 2013-14 and 2015-16 seasons, influenza A (H1N1)pdm09 was the predominant strain. In the 2018-19 season, there were two peaks of similar magnitude dominated by influenza A (H1N1) followed by influenza A (H3N2) (7). During the 2019-2020 season, Influenza B predominated early in the season followed by influenza A (H1N1)pdm09 (8). In the 2020-2021 season, there was unusual low flu activity in the United States when both Influenza A ((H1N1)pdm09)and (H3N2)) and influenza B cocirculated (9), and the majority of influenza A viruses were H3N2 (10). In the 2021-2022 season, the majority of positive flu tests reported to the CDC by US Public Health Laboratories were attributed to influenza A (H3N2) (11).^

^‡The 2019-20 school year is included in pre-COVID-19 seasons and include ILI-SCs through epidemiological week 11 of 2020, the last week of an ILI-related closure in the school year. While first COVID-19-related school closure occurred during epidemiological week 9 in 2020, widespread COVID-19-related school closures were reported during the subsequent epidemiological weeks 12 and 13 of 2020 (5).^

#### ILI-SC CORRELATION WITH SURVEILLANCE OF MEDICALLY ATTENDED ILI

##### **Table S3. Correlation between influenza-like illness-related school closures and percent of outpatient provider visits for ILI^*^ by influenza season and HHS region accounting for three different permutations of the school winter break timing and duration– United States, August 1, 2011—June 30, 2022**

|  | **Weeks Considered as the Winter School Break^‡^** | | | | | |
| --- | --- | --- | --- | --- | --- | --- |
|  | **Week 52 only** | | **Weeks 51 & 52** | | **Weeks 52 & 1** | |
|  | **Rs (95% CI)** | **P** | **Rs (95% CI)** | **P** | **Rs (95% CI)** | **P** |
| **School year/ Influenza season**^§^ |  |  |  |  |  |  |
| 2011-12, H3N2 | 0.425 (0.082, 0.670) | 0.015 | 0.436 (0.090, 0.681) | 0.013 | 0.380 (0.036, 0.636) | 0.029 |
| 2012-13, H3N2 | 0.659 (0.394, 0.816) | <.001 | 0.645 (0.367, 0.810) | <0.001 | 0.569 (0.272, 0.759) | <0.001 |
| 2013-14, H1N1 | 0.309 (-0.050, 0.590) | 0.086 | 0.330 (-0.033, 0.609) | 0.070 | 0.278 (-0.077, 0.564) | 0.118 |
| 2014-15, H3N2 | 0.596 (0.309, 0.776) | <0.001 | 0.677 (0.421, 0.827) | <0.001 | 0.522 (0.215, 0.727) | 0.001 |
| 2015-16, H1N1 | 0.603 (0.313, 0.783) | <0.001 | 0.624 (0.338, 0.798) | <0.001 | 0.583 (0.291, 0.768) | <0.001 |
| 2016-17, H3N2 | 0.833 (0.674, 0.913) | <0.001 | 0.846 (0.695, 0.922) | <0.001 | 0.801 (0.624, 0.895) | <0.001 |
| 2017-18, H3N2 | 0.836 (0.680, 0.915) | <0.001 | 0.819 (0.647, 0.907) | <0.001 | 0.792 (0.608, 0.890) | <0.001 |
| 2018-19, H1N1/ H3N2 | 0.818 (0.648, 0.905) | <0.001 | 0.805 (0.623, 0.900) | <0.001 | 0.775 (0.581, 0.881) | <0.001 |
| 2019-20, H1N1 |  |  |  |  |  |  |
| Week start date (week #) |  |  |  |  |  |  |
| December 15 (week 51) | 0.643 (0.080, 0.883) | 0.022 | 0.530 (-0.128, 0.850) | 0.095 | 0.643 (080, 0.883) | 0.022 |
| January 26 (week 5) | 0.615 (0.173, 0.840) | 0.040 | 0.571 (0.086, 0.826) | 0.019 | 0.434 (-0.054, 0.743) | 0.072 |
| February 23 (week 9) | 0.755 (0.465, 0.891) | 0.001 | 0.738 (0.423, 0.886) | <.001 | 0.580 (0.196, 0.800) | 0.004 |
| March 8 (week 11) | 0.768 (0.508, 0.893) | <0.001 | 0.763 (0.489, 0.893) | <.001 | 0.602 (0.250, 0.804) | 0.001 |
| March 22 (week 13) | 0.619 (0.285, 0.810) | <0.001 | 0.607 (0.258, 0.809) | 0.001 | 0.485 (0.110, 0.730) | 0.011 |
| 2020-21, low circulation^¶^ | 0.289 (-0.065. 0.572) | 0.103 | 0.324 (-0.033, 0.601) | 0.070 | 0.257 (-0.093, 0.545) | 0.143 |
| 2021-22, H3N2 | 0.489 (0.161, 0.712) | 0.004 | 0.552 (0.237, 0.754) | 0.001 | 0.425 (0.089, 0.667) | 0.013 |
| All seasons, 2011-22 | 0.650 (0.584, 0.707) | <0.001 | 0.658 (0.592, 0.715) | <0.001 | 0.604 (0.533, 0.666) | <0.001 |
| Pre-COVID-19 Years^#^, 2011 –20 | 0.678 (0.609, 0.736) | <0.001 | 0.684 (0.614, 0.742) | <0.001 | 0.598 (0.516, 0.668) | <0.001 |
| COVID-19-affected Years ^**^, 2020–22 | 0.475 (0.258, 0.642) | <0.001 | 0.506 (0.291, 0.667) | <0.001 | 0.445 (0.226, 0.617) | <0.001 |
| **HHS region** |  |  |  |  |  |  |
| HHS 1 | 0.150 (-0.500, 0.684) | 0.668 | 0.150 (-0.500, 0.684) | 0.668 | 0.150 (-0.500, 0.684) | 0.668 |
| HHS 2 | 0.295 (-0.290, 0.708) | 0.314 | 0.295 (-0.290, 0.708) | 0.314 | 0.295 (-0.290, 0.708) | 0.314 |
| HHS 3 | 0.311 (-0.084, 0.614) | 0.115 | 0.311 (-0.084, 0.614) | 0.115 | 0.311 (-0.084, 0.614) | 0.115 |
| HHS 4 | 0.662 (0.516, 0.767) | <0.001 | 0.690 (0.548, 0.790) | <0.001 | 0.662 (0.516, 0.767) | <0.001 |
| HHS 5 | 0.614 (0.453, 0.733) | <0.001 | 0.639 (0.482, 0.753) | <0.001 | 0.614 (0.453, 0.733) | <0.001 |
| HHS 6 | 0.753 (0.619, 0.840) | <0.001 | 0.775 (0.649, 0.856) | <0.001 | 0.753 (0.619, 0.840) | <0.001 |
| HHS 7 | 0.424 (0.151, 0.631) | 0.003 | 0.419 (0.138, 0.631) | 0.004 | 0.424 (0.151, 0.631) | 0.003 |
| HHS 8 | -0.465 (-0.802, 0.135) | 0.111 | -0.457 (-0.810, 0.179) | 0.139 | -0.465 (-0.802, 0.135) | 0.111 |
| HHS 9 | -0.258 (-0.975, 0.940) | 0.792 | -0.258 (-0.975, 0.940) | 0.792 | -0.258 (-0.975, 0.940) | 0.792 |
| HHS 10 | 0.281 (-0.093, 0.579) | 0.133 | 0.225 (-0.159, 0.543) | 0.244 | 0.281 (-0.093, 0.579) | 0.133 |

^*^Percent of outpatient provider visits for ILI per data available from ILI Net (3).

^†^Regions of the United States Department of Health & Human Service (HHS) (4).

^‡^Winter break is understood to be approximately 2 weeks in length; however, the start and end dates vary by school and district. Winter breaks consistently overlap on the last week of the year, which was epidemiological week 52 in all years except 2014 and 2020 when it was epidemiological week 53. In 2014 and 2020, we calculated correlations excluding winter break at epidemiological week 53, weeks 52-53, and weeks 53-1.

^§^ In the 2012-13, 2014-15, 2016-17, and 2017-18 influenza seasons, influenza A (H3N2) was the predominant strain (6). In the 2011-12 influenza season, which was unusually mild, influenza A (H3N2) predominated overall but, influenza A (H1N1)pdm09 and influenza B also widely circulated. In the 2013-14 and 2015-16 seasons, influenza A (H1N1)pdm09 was the predominant strain. In the 2018-19 season, there were two peaks of similar magnitude dominated by influenza A (H1N1) followed by influenza A (H3N2) (7). During the 2019-2020 season, Influenza B predominated early in the season followed by influenza A (H1N1)pdm09 (8). In the 2020-2021 season, there was unusual low flu activity in the United States when both Influenza A ((H1N1)pdm09)and (H3N2)) and influenza B cocirculated (9), and the majority of influenza A viruses were H3N2 (10). In the 2021-2022 season, the majority of positive flu tests reported to the CDC by US Public Health Laboratories were attributed to influenza A (H3N2) (11).

^¶^Cannot be calculated because of unusually low flu activity during 2020-2021 season (11).

^#^The 2019-20 school year is included in pre-COVID-19 seasons and includes ILI-SCs through epidemiological week 11 of 2020, the last week of an ILI-related closure in the school year. While the first COVID-19-related school closure occurred during epidemiological week 9 in 2020, widespread COVID-19-related school closures were reported during the subsequent epidemiological weeks 12 and 13 of 2020 (5).

^**^COVID-19-affected years include influenza seasons from the 2020-21 school year through the 2021-22 school year.

##### **Table S4. Correlation between the publicly announced influenza-like illness-related school closures and the reported laboratory-confirmed influenza-associated hospitalizations by age groups and influenza season accounting for three different permutations of the school winter break timing and duration – 13 States^*^, August 1, 2011—June 30, 2022**

|  | **Weeks Considered as the Winter School Break^†^** | | | | | |
| --- | --- | --- | --- | --- | --- | --- |
|  | **Week 52** | | **Weeks 51 & 52** | | **Weeks 52 & 1** | |
|  | **R (95% CI)** | **P** | **R (95% CI)** | **P** | **R (95% CI)** | **P** |
| **Age groups^‡^** |  |  |  |  |  |  |
| All ages | 0.555 (0.473, 0.626) | <0.001 | 0.555 (0.472, 0.628) | <0.001 | 0.517 (0.432, 0.592) | <0.001 |
| 0-4 years old | 0.569 (0.489, 0.639) | <0.001 | 0.570 (0.489, 0.641) | <0.001 | 0.531 (0.448, 0.604) | <0.001 |
| 5-17 years old | 0.592 (0.515, 0.659) | <0.001 | 0.593 (0.514, 0.661) | <0.001 | 0.563 (0.484, 0.633) | <0.001 |
| 18-49 years old | 0.551 (0.468, 0.623) | <0.001 | 0.553 (0.469, 0.626) | <0.001 | 0.511 (0.426, 0.587) | <0.001 |
| 50-64 years old | 0.537 (0.453, 0.611) | <0.001 | 0.537 (0.451, 0.612) | <0.001 | 0.501 (0.415, 0.578) | <0.001 |
| 65+ years old | 0.541 (0.457, 0.614) | <0.001 | 0.542 (0.457, 0.617) | <0.001 | 0.505 (419, 0.582) | <0.001 |
| **All Ages by season^§^**^¶^ |  |  |  |  |  |  |
| 2011-12, H3N2 | 0.466 (0.112, 0.707) | 0.010 | 0.459 (0.095, 0.706) | 0.013 | 0.472 (0.127, 0.707) | 0.008 |
| 2012-13, H3N2 | 0.564 (0.239, 0.767) | 0.001 | 0.541 (0.201, 0.757) | 0.002 | 0.462 (0.115, 0.701) | 0.009 |
| 2013-14, H1N1 | 0.308 (-0.072, 0.603) | 0.105 | 0.332 (-0.053, 0.624) | 0.084 | 0.279 (-0.095, 0.578) | 0.136 |
| 2014-15, H3N2 | 0.492 (0.152, 0.720) | 0.005 | 0.582 (0.265, 0.778) | <0.001 | 0.412 (0.061, 0.665) | 0.020 |
| 2015-16, H1N1 | 0.549 (0.219, 0.758) | 0.002 | 0.546 (0.207, 0.759) | 0.002 | 0.551 (0.229, 0.757) | 0.001 |
| 2016-17, H3N2 | 0.736 (0.496, 0.865) | <0.001 | 0.743 (0.501, 0.871) | <0.001 | 0.696 (0.439, 0.841) | <0.001 |
| 2017-18, H3N2 | 0.745 (0.511, 0.870) | <0.001 | 0.717 (0.458, 0.856) | <0.001 | 0.671 (0.401, 0.827) | <0.001 |
| 2018-19, H1N1/ H3N2 | 0.686 (0.416, 0.837) | <0.001 | 0.671 (0.386, 0.831) | <0.001 | 0.660 (0.383, 0.820) | <0.001 |
| 2019-20, H1N1 | 0.674 (0.350, 0.846) | <0.001 | 0.670 (0.331, 0.847) | <0.001 | 0.597 (0.243, 0.802) | 0.002 |
| 2020-21, low circulation^#^ | - | - | - | - | - | - |
| 2021-22, H3N2 | 0.009 (-0.359, 0.374) | 0.965 | 0.054 (-0.327, 0.418) | 0.788 | -0.035 (-0.390, 0.330) | 0.857 |
| Pre-COVID-19 Years^#^, 2011 – 20 | 0.597 (0.511, 0.670) | <0.001 | 0.601 (0.514, 0.674) | <0.001 | 0.552 (0.461, 0.630) | <0.001 |
| COVID-19-affected Years^**^, 2020 – 22 | 0.324 (0.074, 0.532) | 0.011 | 0.380 (0.129, 0.580) | 0.003 | 0.331 (0.084, 0.536) | 0.009 |
| **Ages 5-17 by season^§^** |  |  |  |  |  |  |
| 2011-12, H3N2 | 0.458 (0.102, 0.702) | 0.012 | 0.444 (0.077, 0.697) | 0.017 | 0.471 (0.126, 0.707) | 0.008 |
| 2012-13, H3N2 | 0.546 (0.215, 0.756) | 0.002 | 0.512 (0.163, 0.739) | 0.005 | 0.458 (0.109, 0.698) | 0.010 |
| 2013-14, H1N1 | 0.294 (-0.087, 0.593) | 0.123 | 0.332 (-0.053, 0.624) | 0.084 | 0.259 (-0.116, 0.563) | 0.169 |
| 2014-15, H3N2 | 0.434 (0.080, 0.683) | 0.016 | 0.518 (0.178, 0.739) | 0.003 | 0.376 (0.018, 0.641) | 0.037 |
| 2015-16, H1N1 | 0.521 (0.182, 0.741) | 0.003 | 0.513 (0.164, 0.740) | 0.005 | 0.528 (0.198, 0.742) | 0.002 |
| 2016-17, H3N2 | 0.862 (0.718, 0.932) | <0.001 | 0.873 (0.733, 0.938) | <0.001 | 0.852 (0.703, 0.926) | <0.001 |
| 2017-18, H3N2 | 0.790 (0.587, 0.894) | <0.001 | 0.784 (0.570, 0.892) | <0.001 | 0.743 (0.513, 0.867) | <0.001 |
| 2018-19, H1N1/ H3N2 | 0.793 (0.591, 0.895) | <0.001 | 0.799 (0.598, 0.900) | <0.001 | 0.768 (0.555, 0.881) | <0.001 |
| 2019-20, H1N1 | 0.773 (0.517, 0.896) | <0.001 | 0.752 (0.470, 0.887) | <0.001 | 0.639 (0.305, 0.824) | <0.001 |
| 2020-21, low circulation^¶^ | - | - | - | - | - | - |
| 2021-22, H3N2 | -0.075 (-0.429, 0.301) | 0.702 | -0.053 (-0.417, 0.328) | 0.793 | -0.095 (-0.439, 0.276) | 0.621 |
| Pre-COVID-19 Years ^**^, 2011 –20 | 0.649 (0.571, 0.714) | <0.001 | 0.653 (0.574, 0.718) | <0.001 | 0.614 (0.533, 0.684) | <0.001 |
| COVID-19-affected Years^††^, 2020 – 22 | 0.223 (-0.35, 0.450) | 0.087 | 0.262 (-0.001, 0.487) | 0.049 | 0.239 (-0.026, 0.453) | 0.076 |

^*^Data on laboratory-confirmed influenza-associated hospitalizations were obtained from FluSurv-NET (12), and were available from the following states: CA, CO, CT, GA, MD, MN, NM, NY, OR, TN, MI, OH, and UT.

^†^School winter break was excluded from the analysis. Winter break is understood to be approximately 2 weeks in length. While the start and end dates vary by school and district, winter breaks consistently overlap on the last week of the year, which was epidemiological week 52 in all years except 2014 and 2020 when it was epidemiological week 53. In 2014 and 2020, we calculated correlations excluding winter break at epidemiological week 53, weeks 52-53, and weeks 53-1.

^‡^Age groups are defined by the data source (FluSurv-NET (12)).

^§^Influenza seasons are epidemiological weeks 40 through 17 (as reported by FluSurv-NET (12)) and shown with the dominant strain of each season.

^¶^In the 2012-13, 2014-15, 2016-17, and 2017-18 influenza seasons, influenza A (H3N2) was the predominant strain (6). In the 2011-12 influenza season, which was unusually mild, influenza A (H3N2) predominated overall but, influenza A (H1N1)pdm09 and influenza B also widely circulated. In the 2013-14 and 2015-16 seasons, influenza A (H1N1)pdm09 was the predominant strain. In the 2018-19 season, there were two peaks of similar magnitude dominated by influenza A (H1N1) followed by influenza A (H3N2) (7). During the 2019-2020 season, Influenza B predominated early in the season followed by influenza A (H1N1)pdm09 (8). In the 2020-2021 season, there was unusual low flu activity in the United States when both Influenza A ((H1N1)pdm09)and (H3N2)) and influenza B cocirculated (9), and the majority of influenza A viruses were H3N2 (10). In the 2021-2022 season, the majority of positive flu tests reported to the CDC by US Public Health Laboratories were attributed to influenza A (H3N2) (11).

^#^Cannnot be calculated because of the unavailability of the weekly hospitalization data for the 2020-2021 season (9).

^**^The 2019-20 school year is included in pre-COVID-19 seasons and includes ILI-SCs through epidemiological week 11 of 2020, the last week of an ILI-related closure in the school year. The first COVID-19-related school closure occurred during epidemiological week 9 in 2020, however widespread COVID-19-related school closures were reported during the subsequent epidemiological weeks 12 and 13 of 2020 (5).

^††^COVID-19-affected years include influenza seasons from the 2020-21 school year through the 2021-22 school year.

Note: Spearman rank correlations were used to evaluate the relationship between Influenza/ILI-related school closures and confirmed laboratory-confirmed influenza-associated hospitalizations during influenza seasons (epidemiological weeks 40 through 17).

##### ILI CLOSURES IN CONTEXT OF OTHER DISEASE-RELATED CLOSURES

Prior to the COVID-19 pandemic, ILI-SCs in our study accounted for 22-87% of annual disease-related SCs and were proportionally highest in the two high severity H3N2-dominated seasons of 2014-2015 and 2017-2018 (Table S6, Fig S2). ILI-SCs accounted for 62% of reactive disease-related reactive closures in 2019-2020, but these all occurred before March 11, 2020 (i.e. before ILI dramatically declined nationwide after the preemptive closures implemented as a COVID countermeasure). Proportionally fewer ILI-SCs occurred in the two school years following the pandemic. These two most recent years also demonstrated a notable decline of SCs related with other diseases (non-ILI/ non-COVID, including gastrointestinal that at times dominated pre-COVID era disease-related SCs, e.g. 2013-2014) and dominance of COVID-19 reactive SCs, including combo announcements of both ILI and COVID.

See Table S5 and Figure S2.

##### **Table S5. Publicly announced, reactive* school closures associated with illness, by school year – United States, August 1, 2011—June 30, 2022**

|  | **School year** | | | | | | | | | | | | | **Total^††^, n (%)** |
| --- | --- | --- | --- | --- | --- | --- | --- | --- | --- | --- | --- | --- | --- | --- |
|  | **Pre-COVID-19 Years^††^** | | | | | | | | | | **COVID-19-affected Years^††^** | | |  |
|  | **2011-12** | **2012-13** | **2013-14** | **2014-15** | **2015-16** | **2016-17** | **2017-18** | **2018-19** | **2019-20^*^** | **Subtotal** | **2020-21** | **2021-22** | **Subtotal** |  |
| Number of school closure events, n (%) | 65 (0.5) | 143 (1.0) | 30 (0.2) | 165 (1.2) | 66 (0.5) | 387 (2.8) | 575 (4.1) | 628 | 1,057 (7.5) | 3,116 | 6,326 (44.9) | 4,646 (33.0) | 10,972 | 14,088 |
| By illness type: |  |  |  |  |  |  |  |  |  |  |  |  |  |  |
| ILI | 33 (50.8) | 104 (72.7) | 5 (16.7) | 112 (67.9) | 20 (30.3) | 231 (59.7) | 458 (79.7) | 429 (68.3) | 627 (59.3) | 2,019 (64.8) | 0 | 9 (0.2) | 9 (0.1) | 2,028 (14.4) |
| Both ILI and COVID | 0 | 0 | 0 | 0 | 0 | 0 | 0 | 0 | 0 | 0 | 5 (0.1) | 44 (1.0) | 49 (0.5) | 49 (0.4) |
| COVID | 0 | 0 | 0 | 0 | 0 | 0 | 0 | 0 | 256 (24.2) | 256 (8.2) | 6,321 (99.9) | 4,520 (97.3) | 10,841 (98.8) | 11,097 (78.8) |
| Gastro-intestinal illness^†^ | 23 (35.4) | 17 (11.9) | 15 (50.0) | 23 (13.9) | 30 (45.5) | 32 (8.3) | 16 (2.8) | 23 (3.7) | 23 (2.2) | 202 (6.5) | 0 | 8 (0.2) | 8 (0.1) | 210 (1.5) |
| Meningitis^‡^ | 1 (1.5) | 2 (1.4) | 1 (3.3) | 2 (1.2) | 7 (10.6) | 1 (0.3) | 4 (0.7) | 9 (1.4) | 2 (0.2) | 29 (0.9) | 0 | 0 | 0 | 29 (0.2) |
| Other respiratory illness^§^ | 1 (1.5) | 1 (0.1) | 3 (10.0) | 3 (1.8) | 2 (3.0) | 8 (2.1) | 1 (0.2) | 19 (3.0) | 5 (0.5) | 43 (1.4) | 0 | 1 (0.0) | 1 (0.0) | 44 (0.3) |
| Other illnesses^¶^ | 1 (1.5) | 1 (0.1) | 0 | 5 (3.0) | 1 (1.5) | 0 | 3 (0.5) | 6 (1.0) | 1 (0.1) | 18 (0.6) | 0 | 0 | 0 | 18 (0.1) |
| Unknown**^#^** | 6 (9.2) | 18 (12.6) | 6 (20.0) | 20 (12.1) | 6 (9.1) | 115 (29.7) | 93 (16.2) | 142 (22.6) | 143 (13.5) | 549 (17.6) | 0 | 64 (1.4) | 64 (0.6) | 613 (4.4) |
| Estimated number of schools closed^**^, n (%) | 169 (0.3) | 637 (1.1) | 51 (0.1) | 407 (0.7) | 117 (0.2) | 1,927 (3.3) | 2,271 (3.9) | 2,551 (4.4) | 4,688 (8.1) | 12,818 | 19,273 (33.2) | 25,924 (44.7) | 45,197 | 58,015 |
| By illness type: |  |  |  |  |  |  |  |  |  |  |  |  |  |  |
| ILI | 103 (61.0) | 383 (60.1) | 11 (21.6) | 308 (75.7) | 40 (34.2) | 1,292 (67.1) | 1,966 (86.6) | 1,912 (75.0) | 2,886 (61.6) | 8,901 (69.4) | 0 | 24 (0.1) | 24 (0.1) | 8,925 (15.4) |
| Both ILI and COVID | 0 | 0 | 0 | 0 | 0 | 0 | 0 | 0 | 0 | 0 | 11 (0.1) | 200 (0.8) | 211 (0.5) | 211 (0.4) |
| COVID | 0 | 0 | 0 | 0 | 0 | 0 | 0 | 0 | 1,185 (25.3) | 1,185 (9.2) | 19,262 (99.9) | 25,479 (98.3) | 44,741 (99.00 | 45,926 (79.2) |
| Gastro-intestinal illness^†^ | 53 (31.4) | 31 (4.9) | 17 (33.3) | 46 (11.3) | 36 (30.8) | 63 (14.7) | 50 (2.2) | 35 (1.4) | 90 (1.9) | 421 (3.3) | 0 | 8 (0.0) | 8 (0.0) | 429 (0.7) |
| Meningitis^‡^ | 1 (0.6) | 3 (0.5) | 1 (2.0) | 3 (6.4) | 19 (16.2) | 1 (2.3) | 4 (0.2) | 11 (0.4) | 4 (0.1) | 47 (0.4) | 0 | 0 | 0 | 47 (0.1) |
| Other respiratory illness^§^ | 1 (0.6) | 1 (0.2) | 12 (23.5) | 5 (1.2) | 5 (4.3) | 16 (0.8) | 1 (0.0) | 22 (0.9) | 5 (0.1) | 68 (0.5) | 0 | 6 (0.0) | 6 (0.0) | 74 (0.1) |
| Other illnesses^¶^ | 1 (0.6) | 5 (0.8) | 0 | 5 (1.2) | 3 (2.6) | 0 | 3 (0.1) | 9 (0.4) | 1 (0.0) | 27 (0.2) | 0 | 0 | 0 | 27 (0.1) |
| Unknown**^#^** | 10 (5.9) | 214 (9.0) | 10 (19.6) | 40 (9.8) | 14 (12.0) | 555 (28.8) | 247 (10.9) | 562 (23.7) | 517 (11.0) | 2.169 (16.9) | 0 | 207 (0.8) | 207 (0.5) | 2,376 (4.1) |

^*^Reactive illness-related closures include only those in response to local transmission of disease. Therefore, this table includes early COVID-19-related closures in response to local transmission, while the pre-emptive COVID-19-related school closures (i.e., those initiated nationwide at the state level in March 2020) are excluded from this table. Majority of the reactive closures in 2019-2020 school year were recorded before March 12, 2020, when the first statewide closure was announced (5).

^†^Gastro-intestinal illness: Stomach flu (due to norovirus or other unspecified gastrointestinal virus), E.coli, shigella, and food/water borne diseases.

^‡^Meningitis includes both bacterial and viral meningitis.

^§^Other reported respiratory illnesses include strep-throat, measles, mumps, pertussis, and Legionnaires' disease.

^¶^Other illnesses-related events : Ebola - 5, hepatitis A - 4, scabies - 4, MRSA - 3, hand, foot, and mouth disease - 1, and varicella - 1. Ebola closures were recorded during week 42 of 2014 in OH and TX, and the reason stated in the announcement was “precaution for staff/students exposed to Ebola”.

**^#^**Unknown or unspecified illness includes unspecified communicable illness, symptoms of high fever/vomiting/headache/stomachache without specifying the illness, dizziness among students.

^**^Schools were counted once for each time they were part of a school closure event at either the district-level or school level.

^††^Percentages may not add up to 100%, as they are rounded to the nearest percent.

##### **Figure S2. Estimated number of school closures associated with illness^*^ (N=58,015) and percent of outpatient provider visits for ILI^†^ by epidemiologic week – United States, August 1, 2011—June 30, 2022**

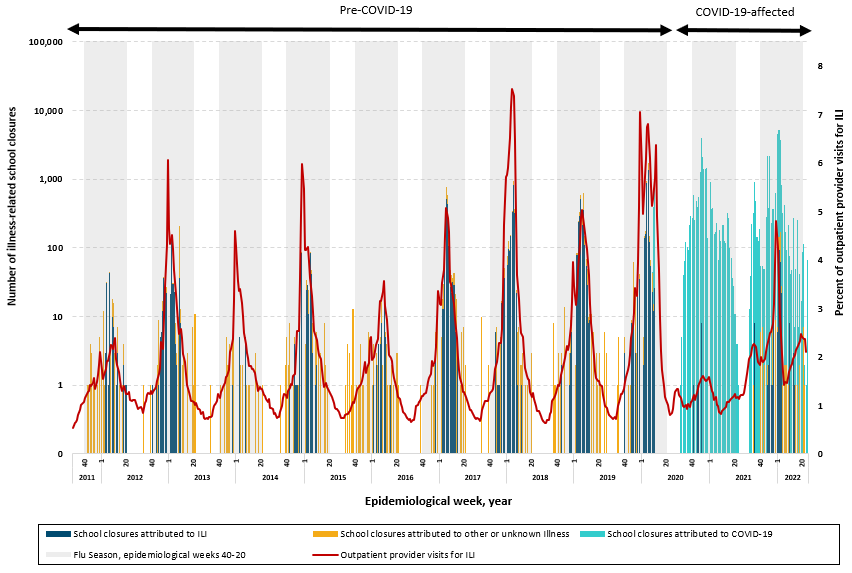

^*^School closures attributed were attributed to three illness categories, [1] ILI which includes both influenza and ILI, [2] COVID-19, [3] unknown or other illness, which includes other respiratory diseases (measles, mumps, pertussis, strep-throat etc.), gastrointestinal diseases, meningitis, and unknown or unspecified illnesses.

^†^Percent of outpatient provider visits for ILI per data available from ILI Net (3).

Note: In the 2012-13, 2014-15, 2016-17, and 2017-18 influenza seasons, influenza A (H3N2) was the predominant strain (6). In the 2011-12 influenza season, which was unusually mild, influenza A (H3N2) predominated overall but, influenza A (H1N1)pdm09 and influenza B also widely circulated. In the 2013-14 and 2015-16 seasons, influenza A (H1N1)pdm09 was the predominant strain. In the 2018-19 season, there were two peaks of similar magnitude dominated by influenza A (H1N1) followed by influenza A (H3N2) (7). During the 2019-2020 season, Influenza B predominated early in the season followed by influenza A (H1N1)pdm09 (8). In the 2020-2021 season, there was unusual low flu activity in the United States when both Influenza A ((H1N1)pdm09)and (H3N2)) and influenza B cocirculated (9), and the majority of influenza A viruses were H3N2 (10). In the 2021-2022 season, the majority of positive flu tests reported to the CDC by US Public Health Laboratories were attributed to influenza A (H3N2) (11).

#### REGRESSION ANALYSIS

During the pre-COVID-19 period, schools located in rural areas had significantly higher odds of experiencing ILI-SCs compared with schools in cities (aOR 8·78, 95% CI 7·93-9·72), followed by towns (town vs city, aOR 6.96, 95% CI 6·25-7·76) (Table S6). Schools with a higher percentage of students enrolled in the federal free or reduced-price lunch program had significant odds to close schools (with 10% increase of enrollment, aOR 1·15, 95% CI 1·14-1·17). Schools with more than the median to upper quartile of student-teacher ratios had significantly increased odds to close schools (aOR 1·77, 95% CI 1·64-1·91) when compared with schools in the lower quartile of student-teacher ratio. However, other school characteristics explored in our analyses such as school type were not significantly associated with ILI-SC occurrence.

During the COVID-19-affected school years, regression analysis led to fewer significant results than in the previous nine school years (Table S6). Schools located in towns had significantly higher odds of experiencing ILI-SCs compared with schools in cities [(aOR 10.86, 95% CI 6·22-18.97)], followed by schools in rural settings [(aOR 5·97, 95% CI 3·42-10·41)]. Only schools with more than the lower quartile to median of student-teacher ratio experienced significant odds to close schools (aOR 1.65, 95% CI: 1.16-2.37) when compared with schools in the lower quartile of student-teacher ratio. The percentage of students enrolled in free/reduced-price lunch were not significantly associated with ILI-SC occurrence.

See Table S6.

##### **Table S6. Selected characteristics of influenza-like illness-related school closures — United States, August 1, 2011—June 30, 2022**

|  | **Pre-COVID-19 Years^*^ (2011-12–2019-20)** | | | | **COVID-19-affected Years^†^ (2020-21–2021-22)** | | | | **All seasons (2011-12–2021-22)** | | | |
| --- | --- | --- | --- | --- | --- | --- | --- | --- | --- | --- | --- | --- |
|  | **Unadjusted OR (95% CI)** | **P** | **Adjusted OR (95% CI)** | **P** | **Unadjusted OR (95% CI)** | **P** | **Adjusted OR**  **(95% CI)** | **P** | **Unadjusted OR (95% CI)** | **P** | **Adjusted OR (95% CI)** | **P** |
| **Urbanicity** |  |  |  |  |  |  |  |  |  |  |  |  |
| City | Ref. |  | Ref. |  | Ref. |  | Ref. |  | Ref. |  | Ref. |  |
| Rural | 4.01 (3.76, 4.29) | <0.001 | 8.78 (7.93, 9.72) | <0.001 | 3.94 (2.54, 6.10) | <0.001 | 5.97 (3.42, 10.41) | <0.001 | 4.02 (3.77, 4.29) | <0.001 | 8.69 (7.87, 9.61) | <0.001 |
| Town | 3.68 (3.42, 3.96) | <0.001 | 6.96 (6.25, 7.76) | <0.001 | 6.75 (4.30, 10.60) | <0.001 | 10.86 (6.22, 18.97) | <0.001 | 3.77 (3.50, 4.05) | <0.001 | 7.11 (6.40, 7.91) | <0.001 |
| Suburban | 0.69 (0.63, 0.75) | <0.001 | 1.04 (0.91, 1.19) | 0.564 | 1.09 (0.64, 1.84) | 0.761 | 0.97 (0.48, 1.98) | 0.940 | 0.70 (0.64, 0.76) | <0.001 | 1.03 (0.90, 1.18) | 0.664 |
| **Student-teacher ratio^‡^** |  |  |  |  |  |  |  |  |  |  |  |  |
| <=Q1 (<=13.37) | Ref. |  | Ref. |  | Ref. |  | Ref. |  | Ref. |  | Ref. |  |
| >Q1 – Q2 (>13.37 – 15.27) | 1.39 (1.31, 1.48) | <0.001 | 1.47 (1.36, 1.59) | <0.001 | 1.66 (1.19, 2.31) | 0.003 | 1.65 (1.16, 2.37) | 0.006 | 1.46 (1.37, 1.55) | <0.001 | 1.52 (1.41, 1.64) | <0.001 |
| >Q2 – Q3 (>15.27 – 17.12) | 1.52 (1.43, 1.61) | <0.001 | 1.77 (1.64, 1.91) | <0.001 | 1.47 (1.01, 2.15) | 0.043 | 1.40 (0.92, 2.13) | 0.114 | 1.63 (1.54, 1.73) | <0.001 | 1.86 (1.72, 2.00) | <0.001 |
| >Q3 (>17.12) | 0.86 (0.81, 0.91) | <0.001 | 1.34 (1.24, 1.45) | <0.001 | 0.84 (0.58, 1.22) | 0.363 | 1.14 (0.77, 1.70) | 0.519 | 0.92 (0.87, 0.98) | 0.007 | 1.41 (1.31, 1.51) | <0.001 |
| **With each 10% increase of students eligible for free/reduced lunch program** | 1.11 (1.10, 1.12) | <0.001 | 1.15 (1.14, 1.17) | <0.001 | 1.01 (0.96, 1.07) | 0.616 | 1.02 (0.96, 1.08) | 0.560 | 1.10 (1.09, 1.11) | <0.001 | 1.14 (1.13, 1.16) | <0.001 |

^*^Pre-COVID-19 years include influenza seasons from the 2011-12 school year, through epidemiological week 11 of 2020 (in the 2019-20 school year), the last week of an ILI-related closure in the school year.

The first COVID-19-related school closure occurred Feb 27 during epidemiological week 9 in 2020(5).

^†^COVID-19-affected years include influenza seasons from the 2020-21 school year through the 2021-22 school year.

**^‡^** Q1=Lower quartile, Q2=Median, Q3=upper quartile.

^§^Private schools were excluded from this analysis (2); the data is available for public schools only (1).

#### REPEAT CLOSURES FOR ILI

Repeat closures for ILI of the same schools were noted in the data, occurring both within and across school years. Multiple ILI-SCs per school within an individual school year were highest in the three years leading up to the COVID-19 pandemic, and non-existent in the COVID-affected period (Table S7). The majority of the repeat closures within the same school year exhibited a steady annual increase in the pre-pandemic period and were concentrated in HHS 4, occurring in H3N2 dominated seasons, and limited to rural areas (Table S8). Repeat closure of schools for ILI across school years were experienced by more than a third of affected schools and demonstrated a similar pattern with the majority occurring in HHS 4 and rural areas (Table S9).

See Tables S7, S8, and S9.

##### **Table S7. Recurrence of influenza-like illness-related school closures within each school year among unique schools^*^, United States, August 1, 2011 – June 30, 2022**

| School year | Estimated number of closed schools^†^ (n=9,136) ^§^ | Affected schools by number of unique ILI-related closures^§^: | | |
| --- | --- | --- | --- | --- |
|  |  | 1X (7,154) | 2X (n=892) | 3X (n=66) |
| 2011-12 | 103 (1.1) | 89 (92.7) | 7 (7.3) | 0 |
| 2012-13 | 383 (4.2) | 313 (90.5) | 29 (8.4) | 4 (1.2) |
| 2013-14 | 11 (0.1) | 11 (100) | 0 | 0 |
| 2014-15 | 308 (3.4) | 296 (98.0) | 6 (2.0) | 0 |
| 2015-16 | 40 (0.4) | 32 (88.9) | 4 (11.1) | 0 |
| 2016-17 | 1,292 (14.1) | 1,048 (90.0) | 107 (9.2) | 10 (0.9) |
| 2017-18 | 1,966 (21.5) | 1,525 (87.4) | 219 (12.6) | 1 (0.1) |
| 2018-19 | 1,912 (20.9) | 1,444 (86.1) | 234 (13.9) | 0 |
| 2019-20^‡^ | 2,886 (31.6) | 2,161 (86.5) | 286 (11.4) | 51 (2.0) |
| 2020-21 | 11 (0.1) | 11 (100) | 0 | 0 |
| 2021-22 | 224 (2.5) | 224 (100) | 0 | 0 |

^*^Unique school: each school experiencing closure was counted only once per year, regardless of the number of times it closed.

^†^Numbers of schools in district-level closure events were estimated based on the number of K-12 schools in each affected school district per data available from the National Center for Education Statistics (1)^.^ Schools were counted once for each time they were part of a school closure event at either the district-level or school level.

^‡^The last ILI-related closure for this school year was documented on March 11 (week 11), the day before the announcement of the first statewide closure due to COVID-19 pandemic (5).

^§^Percentages may not add up to 100%, as they are rounded to the nearest percent.

##### **Table S8. Characteristics of schools with repeat closures due to influenza-like illness by school year -- United States, August 1, 2011—June 30, 2022**

|  | **School year** | | | | | | | | | | |
| --- | --- | --- | --- | --- | --- | --- | --- | --- | --- | --- | --- |
|  | **Pre-COVID-19 Years (n=958)** | | | | | | | | | **COVID-19-affected Years (n=0)** | |
|  | **2011-12** | **2012-13** | **2013-14** | **2014-15** | **2015-16** | **2016-17** | **2017-18** | **2018-19** | **2019-20^*^** | **2020-21** | **2021-22** |
|  | n=7 (0.7) | n=33 (3.4) | n=0 | n=6 (0.6) | n=4 (0.4) | n=117 (12.2) | n=220 (23.0) | n=234 (24.4) | n=337 (35.2) | n=0 | n=0 |
| HHS region for the affected schools^†^, n (%) | HHS 4, 7 (100.0) | HHS 4, 27 (81.8)  HHS 5, 1 (3.0)  HHS 6, 5 (15.2) |  | HHS 4, 6 (100.0) | HHS 5, 3 (75.0)  HHS 10, 1 (25.0) | HHS 4, 108 (82.3)  HHS 5, 2 (1.7)  HHS 6, 6 (5.1)  HHS 7, 1 (0.9) | HHS 4, 191 (86.8)  HHS 5, 12 (5.5)  HHS 6, 10 (4.6)  HHS 7, 4 (1.8)  HHS 10, 3 (1.4) | HHS 3, 2 (0.9)  HHS 4, 202 (86.3)  HHS 5, 3 (1.3)  HHS 6, 17 (7.3)  HHS 7, 6 (2.6)  HHS 10, 4 (1.7) | HHS 4, 295 (87.5)  HHS 5, 18 (5.3)  HHS 6, 23 (6.8)  HHS 10, 1 (0.3) |  |  |
| H3N2-dominant Influenza season^‡^ (Yes/No) | Yes | Yes | No | Yes | No | Yes | Yes | Yes | No | Low circulation | Yes |
| Urbanicity of the affected schools, n (%) | Rural, 7 (100.0) | Rural, 29 (87.9)  Town, 3 (9.1)  City, 1 (3.0) |  | Rural, 1 (16.7)  Town, 5 (83.3) | Rural, 3 (75.0)  City, 1 (25.0) | Rural, 64 (54.7)  Town, 40 (34.2)  Suburban, 13 (11.1) | Rural, 96 (43.6)  Town, 32 (14.6)  Suburban, 34 (15.5)  City, 58 (26.4) | Rural, 100 (42.7)  Town, 38 (16.2)  Suburban, 43 (18.4)  City, 52 (22.2)  Not specified, 1 (0.4) | Rural, 205 (60.8)  Town, 106 (31.5)  Suburban, 14 (4.2)  City, 11 (3.3)  Not specified, 1 (0.3) |  |  |

^*^The 2019-20 school year is included in pre-COVID-19 seasons and includes ILI-SCs through epidemiological week 11 of 2020, the last week an ILI-related closure was reported in the school year. While the first COVID-19-related school closure occurred during epidemiological week 9 in 2020, widespread COVID-19-related schools closures were reported during the subsequent epidemiological weeks 12 and 13 of 2020 (5).

^†^Regions of the United States Department of Health & Human Service (HHS) (4).

^‡^ In the 2012-13, 2014-15, 2016-17, and 2017-18 influenza seasons, influenza A (H3N2) was the predominant strain (6). In the 2011-12 influenza season, which was unusually mild, influenza A (H3N2) predominated overall but, influenza A (H1N1)pdm09 and influenza B also widely circulated. In the 2013-14 and 2015-16 seasons, influenza A (H1N1)pdm09 was the predominant strain. In the 2018-19 season, there were two peaks of similar magnitude dominated by influenza A (H1N1) followed by influenza A (H3N2) (7). During the 2019-2020 season, Influenza B predominated early in the season followed by influenza A (H1N1)pdm09 (8). In the 2020-2021 season, there was unusual low flu activity in the United States when both Influenza A ((H1N1)pdm09)and (H3N2)) and influenza B cocirculated (9), and the majority of influenza A viruses were H3N2 (10). In the 2021-2022 season, the majority of positive flu tests reported to the CDC by US Public Health Laboratories were attributed to influenza A (H3N2) (11).

##### **Table S9. Recurrence of influenza-like illness (ILI)-related school closures among unique schools^*^– United States, August 1, 2011 – June 30, 2022**

|  | **School years** | | | | | | | |
| --- | --- | --- | --- | --- | --- | --- | --- | --- |
|  | **Pre-COVID-19 Years**^§^ | | | | | | **COVID-19-affected Years**^§^ | |
| **Selected characteristics** | **Unique schools, n (%)** | **Number of schools experiencing single ILI-related closure** | **Number of schools with multiple ILI-related closures** | | | | **Unique schools, n (%)** | **Number of schools experiencing single ILI-related closure** |
|  |  |  | Schools with 2 or more ILI-SCs | 2X | 3-5X | 6-11X |  |  |
| Total^†^ | 4,824 | 2,979 (61.8) | 1,845 (38.2) | 815 (16.9) | 805 (16.7) | 225 (4.7) | 235 | 235 (100.0) |
| Urbanicity |  |  |  |  |  |  |  |  |
| City | 764 (15.8) | 568 (19.1 | 196 (10.6) | 71 (8.7) | 77 (9.6) | 48 (21.3) | 25 (10.6) | 25 (10.6) |
| Suburban | 481 (10.0) | 272 (9.1) | 209 (11.3) | 85 (10.4) | 82 (10.2) | 42 (18.7) | 31 (13.2) | 31 (13.2) |
| Town | 1,043 (21.6) | 586 (19.7) | 457 (24.8) | 211 (25.9) | 209 (26.0) | 37 (16.4) | 77 (32.8) | 77 (32.8) |
| Rural | 2,517 (52.2) | 1,537 (51.6) | 980 (53.1) | 445 (54.6) | 437 (54.3) | 98 (43.6) | 101 (43.0) | 101 (43.0) |
| Not specified | 19 (0.4) | 16 (0.5) | 3 (0.2) | 3 (0.4) | 0 | 0 | 1 (0.4) | 1 (0.4) |
| HHS Region^‡^ |  |  |  |  |  |  |  |  |
| HHS 1 | 14 (0.3) | 14 (0.5) | 0 | 0 | 0 | 0 | 7 (3.0) | 7 (3.0) |
| HHS 2 | 39 (0.8) | 35 (1.2) | 4 (0.2) | 4 (0.5) | 0 | 0 | 4 (1.7) | 4 (1.7) |
| HHS 3 | 172 (3.6) | 155 (5.2) | 17 (0.9) | 17 (2.1) | 0 | 0 | 0 | 0 |
| HHS 4 | 2,372 (49.2) | 897 (30.1) | 1,475 (80.0) | 489 (60.0) | 761 (94.5) | 225 (100.0) | 26 (11.1) | 26 (11.1) |
| HHS 5 | 819 (17.0) | 714 (24.0) | 105 (5.7) | 90 (11.0) | 15 (1.9) | 0 | 49 (20.9) | 49 (20.9) |
| HHS 6 | 1,048 (21.7) | 844 (28.3) | 204 (11.1) | 176 (21.6) | 28 (3.5) | 0 | 86 (36.6) | 86 (36.6) |
| HHS 7 | 200 (4.2) | 177 (5.9) | 23 (1.3) | 22 (2.7) | 1 (0.1) | 0 | 27 (11.5) | 27 (11.5) |
| HHS 8 | 38 (0.8) | 38 (1.3) | 0 | 0 | 0 | 0 | 0 | 0 |
| HHS 9 | 3 (0.1) | 3 (0.1) | 0 | 0 | 0 | 0 | 11 (4.7) | 11 (4.7) |
| HHS 10 | 119 (2.5) | 102 (3.4) | 17 (0.9) | 17 (2.1) | 0 | 0 | 25 (10.6) | 25 (10.6) |

^*^Unique school: During the study period, each school experiencing closure was counted only once.

^†^Total row presented with row percent, all else reported with column percent.

^‡^Regions of the United States Department of Health & Human Services (HHS) (4).

^§^Percentages may not add up to 100%, as they are rounded to the nearest percent.
